# Characterising grey-white matter relationships in recent-onset psychosis and its association with cognitive function

**DOI:** 10.1101/2024.08.13.24311953

**Authors:** Yoshito Saito, Christos Pantelis, Vanessa Cropley, Liliana Laskaris, Cassandra M. J. Wannan, Warda T. Syeda

**Author notes:** These authors contributed equally: Cassandra M. J. Wannan and Warda T. Syeda.

## Abstract

Individuals with recent-onset psychosis (ROP) present widespread grey matter (GM) reductions and white matter (WM) abnormalities. However, relationships between GM and WM changes and their association with cognitive impairment, a key symptom of ROP, are unclear. Using a multiblock partial least squares correlation (MB-PLS-C) analysis, we examined multivariate GM-WM relationships and their association with cognitive abilities in ROP. We used T1 and diffusion-weighted images from 71 non-affective ROP individuals (age 22.1±3.2) and 71 matched controls. We performed MB-PLS-C between GM thickness and WM fractional anisotropy (FA) and between GM surface area and WM FA to identify multivariate GM-WM patterns and analysed correlations between these patterns and cognitive abilities. MB-PLS-C identified a ‘GM thickness’-‘WM FA’ pattern representing group differences, explaining 12.38% of the variance and associated with frontal and temporal GM regions and seven WM tracts, including the corticospinal tract. MB-PLS-C also identified a ‘GM surface area’-‘WM FA’ pattern showing group differences, explaining 18.92% and related with cingulate, frontal, temporal, and parietal GM regions and 15 WM tracts, including the inferior cerebellar peduncle. The ‘GM thickness’-‘WM FA’ pattern describing group differences was significantly correlated with processing speed in ROP. There was no association between cognition and the ‘GM surface area’-‘WM FA’ pattern. MB-PLS-C identified differential whole-brain GM-WM relationships, indicating a potential signature of brain alterations in ROP. Our findings of a relationship between cognitive function and GM-WM patterns for GM thickness rather than for surface area have implications for our understanding of brain-behaviour relationships neurodevelopmentally in psychosis.

## INTRODUCTION

Widespread grey matter (GM) and white matter (WM) abnormalities are observed in both early psychosis and chronic schizophrenia-spectrum disorders, with the greatest degree of change observed in the first 2-5 years of illness [1–7]. Recent studies suggest that these GM and WM abnormalities may be interrelated, demonstrating that cortical thinning in various stages of schizophrenia-spectrum illness is linked to changes in the adjacent or connected WM and occurs coordinatedly through connected regions [8–13].

While regional changes in GM thickness have been associated with changes in adjacent or connected WM, it is important to consider that many GM regions are not only directly connected but also indirectly connected through key relay points [10,12,13]. Brain network analyses have demonstrated that using only the shortest paths between brain regions would ignore a significant portion of existing WM routes and that indirect routes are considered necessary for brain flexibility [14–16]. Multiple studies indicate GM changes in schizophrenia-spectrum disorder, including first-episode psychosis, occur in a coordinated manner between regions that are structurally or functionally associated [8,9,11]. To fully understand widespread brain alterations in schizophrenia-spectrum disorder, it is therefore important to consider correlated relationships between changes in GM and WM, not only in directly connected regions, but also throughout the entire brain. Given that the most significant GM and WM changes occur 2-5 years after onset, it would be valuable to explore correlated GM and WM changes in recent-onset psychosis [4,17].

Partial least squares correlation (PLS-C) provides one possible approach to examining multivariate whole-brain GM-WM relationships [18,19]. PLS-C is a dimensionality reduction technique to find key latent correlations between two sets of variables, identifying patterns overlapping between groups or representing group differences. Recent studies have utilised PLS-C to investigate underlying relationships between regional brain measures, such as cortical thickness, and behavioural data, including cognitive functioning, in schizophrenia-spectrum disorder [20–24]. However, to date, this method has not been used to examine relationships between GM and WM. PLS-C could identify latent correlation patterns between multiple GM regions and WM tracts, contributing to the understanding of complex whole-brain interactions. Recently, we have developed a novel PLS-C approach, multiblock PLS-C (MB-PLS-C), which incorporates covariates into PLS-C as a separate block and demonstrates the effects of covariates on each latent pattern [24]. MB-PLS-C could possibly identify whole-brain correlated GM-WM changes in schizophrenia-spectrum disorder.

Gaining a better understanding of correlated GM-WM patterns in schizophrenia-spectrum disorder is also important for understanding cognitive impairments, such as processing speed, working memory, and episodic memory, in these disorders [25]. Cognitive impairments are observed from the early stages of psychosis and are associated with functional impairments and poor outcomes [26–29]. However, a recent systematic review indicates that these impairments are largely not correlated with GM abnormalities, except for episodic memory impairment, which has been associated with hippocampal volumes [30]. Furthermore, while a handful of studies have identified relationships between fractional anisotropy of specific WM tracts and cognitive abilities in schizophrenia-spectrum disorder [31–34], other studies, including a large-sample ENIGMA consortium study, did not identify specific WM-cognition relationships [35–39]. However, existing studies have largely evaluated relationships between individual brain structures and specific cognitive abilities. Given that cognitive functions are associated with brain networks and interactions among multiple brain regions [40–42], examining relationships between cognition and whole-brain GM-WM patterns may provide a greater understanding of the neural correlates of cognitive impairment in schizophrenia-spectrum disorder.

In this study, we aimed to 1) identify multivariate and coupled whole-brain GM-WM patterns in recent-onset psychosis (ROP) using MB-PLS-C and 2) examine how these GM-WM patterns are associated with cognitive abilities. We hypothesised that MB-PLS-C between GM and WM would identify two distinct patterns: one that overlaps between healthy controls and ROP individuals and one that shows group differences. We also hypothesised that cognitive impairments across multiple domains would be associated with the GM-WM pattern representing a difference between these groups. Finally, we assessed the anatomical plausibility of these multivariate GM-WM patterns using structural connectivity.

## MATERIALS AND METHODS

### Participants

The study included 71 individuals with recent-onset psychosis (ROP) within five years of their first psychotic episode from the Human Connectome Project for Early Psychosis (HCP-EP) and 71 healthy controls (HCs) from two datasets: HCP-EP and the HCP-Development (HCP-D) [43,44]. The controls were selected from healthy individuals within the HCP-EP and HCP-D using propensity score matching so that the number of participants, sex, and age were matched between the groups [45,46]. Demographic and clinical data are displayed in Table 1. Details of the datasets and propensity score matching are described in the Supplementary Material (see Figure S1 and Table S1).

**Table 1.**
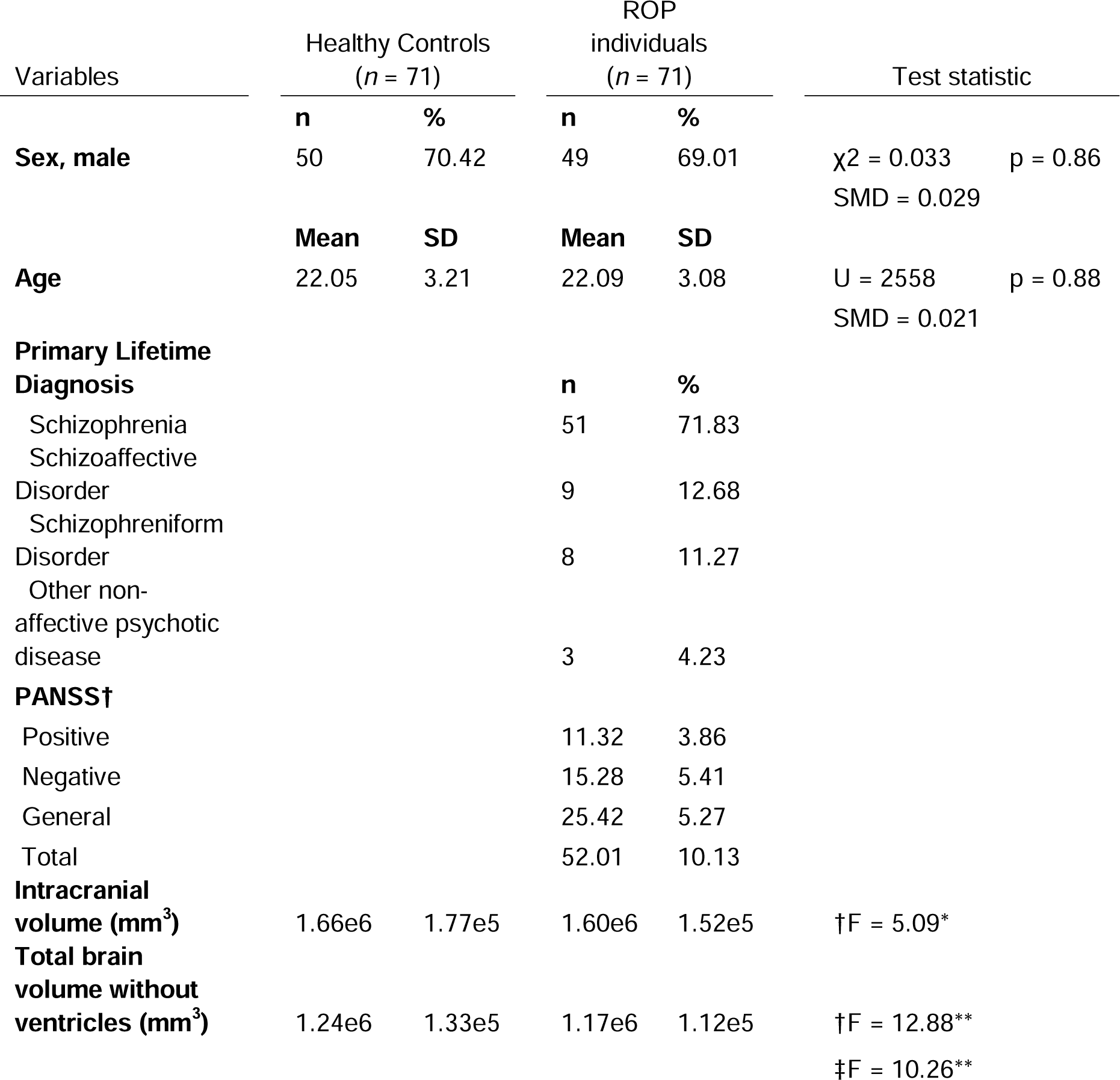
Demographic and clinical data. Key: Standardised mean difference (SMD) ≤ 0.1 evaluates the mean difference of continuous and binary variables between groups. The SMD of ≤ 0.1 indicates the covariate is well-matched between groups. * 0.01≤*p*<0.05, ** *p*<0.01 † the *F*-statistic for analysis of covariance with age and sex as covariate ‡ the *F*-statistic for analysis of covariance with age, sex and intracranial volume as covariate

| Variables | Healthy Controls<br>(*n* = 71) |  | ROP<br>individuals<br>(*n* = 71) |  | Test statistic |  |
| --- | --- | --- | --- | --- | --- | --- |
|  | **n** | **%** | **n** | **%** |  |  |
| **Sex, male** | 50 | 70.42 | 49 | 69.01 | $\chi^2 = 0.033$ | $p = 0.86$ |
|  |  |  |  |  | SMD = 0.029 |  |
|  | **Mean** | **SD** | **Mean** | **SD** |  |  |
| **Age** | 22.05 | 3.21 | 22.09 | 3.08 | $U = 2558$ | $p = 0.88$ |
|  |  |  |  |  | SMD = 0.021 |  |
| **Primary Lifetime**<br>**Diagnosis** |  |  | **n** | **%** |  |  |
| Schizophrenia |  |  | 51 | 71.83 |  |  |
| Schizoaffective<br>Disorder |  |  | 9 | 12.68 |  |  |
| Schizophreniform<br>Disorder |  |  | 8 | 11.27 |  |  |
| Other non-affective psychotic<br>disease |  |  | 3 | 4.23 |  |  |
| **PANSS†** |  |  |  |  |  |  |
| Positive |  |  | 11.32 | 3.86 |  |  |
| Negative |  |  | 15.28 | 5.41 |  |  |
| General |  |  | 25.42 | 5.27 |  |  |
| Total |  |  | 52.01 | 10.13 |  |  |
| **Intracranial**<br>**volume (mm<sup>3</sup>)** | 1.66e6 | 1.77e5 | 1.60e6 | 1.52e5 | †F = 5.09* |  |
| **Total brain**<br>**volume without**<br>**ventricles (mm<sup>3</sup>)** | 1.24e6 | 1.33e5 | 1.17e6 | 1.12e5 | †F = 12.88** |  |
|  |  |  |  |  | ‡F = 10.26** |  |

### Cognitive assessment

In the NIH cognition toolbox (Table S2), this study included analyses of working memory, episodic memory, and processing speed [47–49]. These cognitive functions have shown large effect sizes between patients and controls in past meta-analyses and were identified as three main cognitive categories in schizophrenia [50–52]. Word reading ability, a measure of pre-morbid IQ, was also included [53, 54].

### MRI acquisition and processing

T1- and diffusion-weighted images were used from the HCP databank. T1-weighted images were processed by the HCP consortium, and we processed the diffusion-weighted images with the HCP minimal preprocessing pipeline [55]. Thickness, surface area, and volume of GM regions were calculated using FreeSurfer (v6.0.0) (https://surfer.nmr.mgh.harvard.edu) according to the Desikan-Killiany Atlas [56]. WM parameters, including fractional anisotropy (FA) and mean diffusivity (MD), were computed using tract-based spatial statistics analysis with FSL (v6.0.4) for WM tracts of Johns Hopkins University white matter tract atlas [57]. Harmonisation using ComBat was applied to GM and WM variables to reduce the effect of the scanner and protocol differences [58–60]. The details are provided in the Supplementary Material (see Figures S2-4 and Tables S3, 4).

### Statistical analyses

Demographic, clinical, cognitive, and GM/WM measures were compared between groups using the Mann-Whitney U test, chi-square test, or ANCOVA based on data type and covariates. Correlations were assessed using Pearson’s or Spearman’s correlation coefficient based on the normality of the data.

### Multiblock partial least squares correlation analyses (MB-PLS-C)

This study employed MB-PLS-C to identify multivariate GM-WM patterns. MB-PLS-C identifies latent variables (LVs) that maximise covariance between two multivariate data blocks and between data and covariate blocks [24]. Details of MB-PLS-C are in the Supplementary Material. Briefly, a multiblock correlation matrix between GM and WM variables with their covariates was decomposed into singular vector matrices and a singular value matrix by singular value decomposition. Singular vector matrices represent the relevance of GM regions and WM tracts to respective LVs, referred to as ‘salience’ in this paper.

Permutation tests assessed the significance of the overall pattern and LVs with 10 000 permutations, and the confidence interval for saliences was estimated by bootstrapping with 10 000 iterations. The model generalisability was assessed by cross-validation and an out-of-sample analysis. The details are presented in the Supplementary Material (see Table S5).

### Interpretation of GM/WM saliences

The salience pattern of each LV illustrates the contribution of variables to their corresponding LV. The pattern of GM variables reflects the polarity of the saliences per group. A pattern overlapping between groups is indicated if each group’s salience distribution shows the same direction, while a pattern showing group differences appears if a distribution from each group has a different direction. Saliences of WM variables represent an overall pattern averaged across the groups. The product of GM and WM saliences reflects the strength of a GM-WM pattern, and the polarity of the product suggests the direction of the pattern.

### Further Analysis of MB-PLS-C Results

Group differences in GM patterns were evaluated by the normalised between-group structural salience difference (NSSD) for each LV [24]. To examine the convergence of salience values with increasing sample size, we calculated the absolute difference in each salience between N+1 and N samples and plotted trajectories. Please refer to the Supplementary Material for further details.

### Tractography-based validation of GM-WM patterns derived from MB-PLS-C

To investigate the anatomical plausibility of the GM-WM patterns describing group differences, we performed tractography and calculated the total weight of connections for the GM-WM patterns (see Figure 5 in the results section) [61,62]. We calculated the average weight of streamlines passing the *k*th WM tract from the *i*th to *}*th GM region (1<=*i, }*<=68, 1<=*k*<=44) for each group. We summed the weight of streamlines in the GM-WM pattern. To assess if the connectivity of the GM-WM pattern was significantly high, we generated 10 000 random GM-WM patterns, each consisting of the same number of GM regions and WM tracts as the original pattern. The p-value was determined by the proportion of random GM-WM patterns with a greater total streamline weight than the total in the original GM-WM pattern. More details are in the Supplementary Material.

### Correlation between cognitive abilities and LVs of GM-WM patterns showing group differences

The correlations between cognitive abilities and latent variables of GM-WM patterns, with GM saliences in different directions between the groups, were evaluated in ROP individuals and controls separately. We focused on the latent variables indicating the GM-WM relationship, rather than the relationship with covariates. A group difference in a correlation was analysed by Fisher’s z-test.

### Follow-up analyses

The correlations between latent variables from MB-PLS-C analysis and three PANSS scores (positive, negative, and general psychopathology scales) and antipsychotic doses were evaluated. By using allometric scaling maps, we evaluated the similarity between cortical variations based on typical development and the latent cortical patterns identified through MB-PLS-C analysis [63]. Details of allometric scaling maps are described in the Supplementary Material.

## RESULTS

### Case-control differences in cognition, GM, and WM

Significant between-group differences were observed for all cognitive measures, with poorer performance in ROP individuals (Table S6). Lower GM volume was observed in regions of the parietal and temporal lobes, as well as the nucleus accumbens bilaterally, in ROP individuals (Figure S6). GM thickness showed widespread reduction, primarily in the temporal, parietal, and frontal lobes and insula. No between-group differences were observed in GM surface area, as well as in FA and MD of any WM tracts (Figure S7).

### MB-PLS-C between GM thickness and WM FA

The MB-PLS-C analyses between GM thickness and WM FA were significant (omnibus test *p*<0.0005). The input multiblock cross-correlation matrix was decomposed into latent matrices by MB-PLS-C (Figure S8). While LV1 was not significant, LV2 and LV3 were significant and explained 29.30% of sum-of-squares variance (Table S7).

### Second latent variable (LV2)

LV2 accounted for 16.92% of sum-of-squares variance (*p*=0.040) and reflected correlations in controls, with negative correlations observed between increased age and decreased thickness in the covariate blocks (Figure S8).

For GM and WM saliences, the sign (positive or negative) itself does not carry meaning, and their direction has meaning (see “Interpretation of GM/WM saliences” in the Methods section). Almost all significant GM saliences in controls were consistently large and in the same direction, while those in the ROP group were more variable, sparse, and smaller (Figure 1A). The largest GM saliences were observed in frontal and parietal regions in controls, whereas those were observed in cingulate and temporal regions in ROP individuals. The largest positive WM saliences were observed in the bilateral cingulum in cingulate and parahippocampal gyri and left medial lemniscus, with the largest negative saliences in the bilateral cerebral peduncle (Figure 1B).

**Figure 1.**
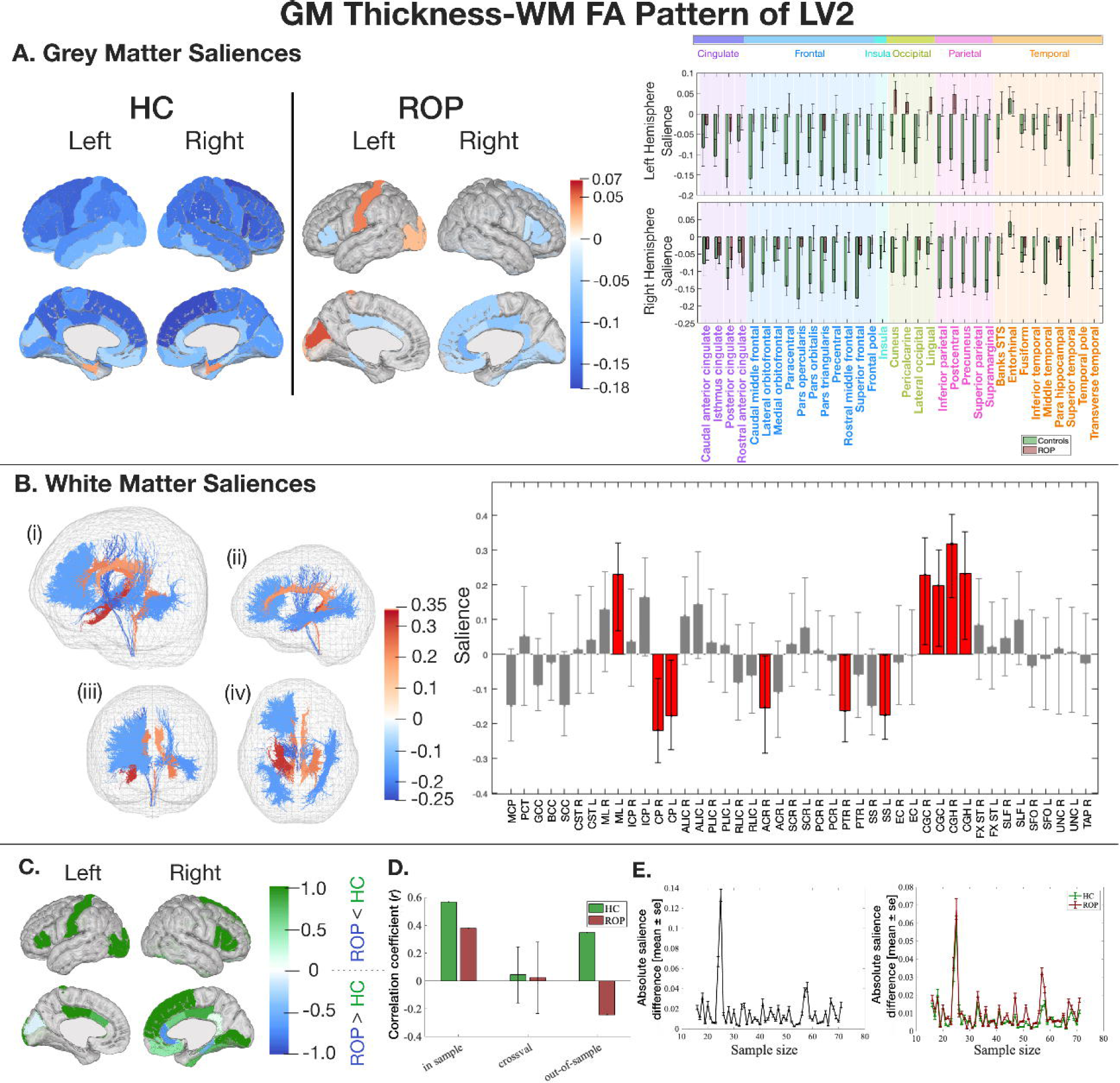
The latent pattern of LV2 between GM thickness and WM FA derived from MB-PLS-C. Key: A) GM salience by group in HC and ROP individuals. GM saliences of HC (green) and ROP (red) with 95% confidence interval (black lines) and color-coded lobe information. B) WM saliences shared between groups. WM tracts represented in tractography are colour-coded based on the salience intensity ((i) overall view (ii) from the left (iii) from the front (iv) from the bottom). The bar plot shows the salience of each WM tract with 95% confidence intervals (black lines). C) Normalised regional salience-differences. A positive score (green) shows a salience is larger in HC, and a negative score (blue) shows a salience is larger in ROP. D) A correlation coefficient between LV2 of GM and WM in the training and the test samples, and a correlation coefficient and its standard deviation from Monte Carlo cross-validation. The cross-validation data did not show a significant correlation, and the out-of-sample results only demonstrated a generalisable correlation for controls. E) The impact of sample size on salience intensity. Figures on the left and right depict differences in WM saliences and GM saliences, respectively. The GM-WM pattern converged with 25 samples from each group. ACR=anterior corona radiata; ALIC=anterior limb of the internal capsule; BCC=body of corpus callosum; CGC=cingulum (cingulate gyrus); CGH=cingulum (hippocampal portion); CP=cerebral peduncle; CR=corona radiata; CST=corticospinal tract; EC=external capsule; FX ST=fornix (cres) / stria terminalis; GCC=genu of corpus callosum; ICP=interior cerebellar peduncle; ML=medial lemniscus; MCP=middle cerebellar peduncle; PCT=pontine crossing tract; PCR=posterior corona radiata; PLIC=posterior limb of the internal capsule; PTR=posterior thalamic radiation; RLIC=retrolenticular part of the internal capsule; SCC=splenium of corpus callosum; SCR=superior corona radiata; SFO=superior fronto-occipital fasciculus; SLF=superior longitudinal fasciculus; SS=sagittal stratum; TAP=tapetum; UNC=uncinate fasciculus.

The LV-specific NSSD metric shows the group differences in GM saliences. The HC group showed the most robust NSSD metrics in the right pars opercularis, superior frontal gyrus, and pars triangularis while the ROP group exhibited those in the right rostral anterior cingulate and parahippocampal gyri and left cuneus (Figure 1C). In the training sample (n=142), both groups showed a positive correlation between latent variables of GM thickness and WM FA with a greater correlation in the HCs (HC: *r*=0.57, p<0.001, ROP: *r*=0.38, *p* =0.001). A significant correlation was not observed in the cross-validation data (n=112 in the training set, n=30 in the test set), and only controls showed a generalisable correlation in the out-of-sample results (n=51) (Figure 1D). GM and WM saliences showed convergence when sample size reached 25 samples per group (Figure 1E).

In summary, LV2 represented a significant pattern derived from the relationship between GM thickness and age in controls, involving frontal and parietal GM regions and WM tracts, including the bilateral cingulum. The salience patterns were stable in this sample size.

### Third latent variable (LV3)

LV3 explained 12.38% of sum-of-squares variance (*p*=0.003), with controls displaying negative correlations between GM thickness and WM FA, whereas ROP individuals demonstrated positive correlations (Figure S8).

A GM salience pattern described group differences, where saliences were positively and strongly mapped onto the ROP group while controls showed smaller negative saliences (Figure 2A). The largest GM saliences were observed in occipital and temporal regions in controls, whereas those were observed in frontal and temporal regions in ROP individuals. The largest negative WM saliences were present in the bilateral anterior limb of the internal capsule, left posterior thalamic radiation and retrolenticular limb of the internal capsule, and right corticospinal tract (Figure 2B).

**Figure 2.**
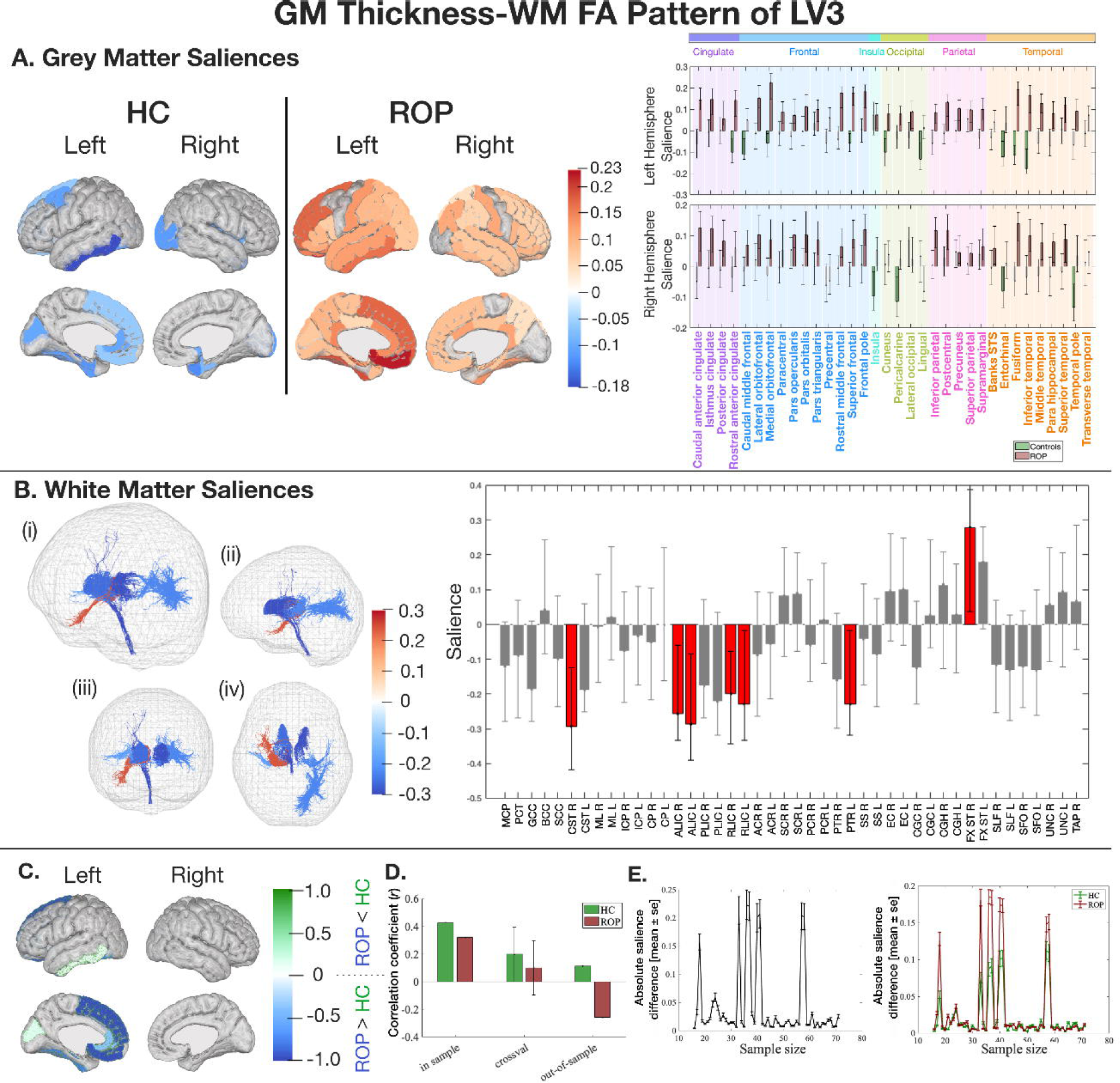
The latent pattern of LV3 between GM thickness and WM FA derived from MB-PLS-C. Key: A) GM salience by group in HC and ROP individuals. GM saliences of HC (green) and ROP (red) with 95% confidence interval (black lines) and color-coded lobe information. B) WM saliences shared between groups. WM tracts represented in tractography are colour-coded based on the salience intensity ((i) overall view (ii) from the left (iii) from the front (iv) from the bottom). The bar plot shows the salience of each WM tract with 95% confidence intervals (black lines). C) Normalised regional salience-differences. A positive score (green) shows a salience is larger in HC, and a negative score (blue) shows a salience is larger in ROP. D) A correlation coefficient between LV3 of GM and WM in the training and the test samples, and a correlation coefficient and its standard deviation from Monte Carlo cross-validation. Only the the cross-validation data from the HC group exhibited a generalised correlation, and the out-of-sample data did not demonstrate one. E) The impact of sample size on salience intensity. Figures on the left and right depict differences in WM saliences and GM saliences, respectively. The GM-WM pattern converged with 60 samples from each group. ACR=anterior corona radiata; ALIC=anterior limb of the internal capsule; BCC=body of corpus callosum; CGC=cingulum (cingulate gyrus); CGH=cingulum (hippocampal portion); CP=cerebral peduncle; CR=corona radiata; CST=corticospinal tract; EC=external capsule; FX ST=fornix (cres) / stria terminalis; GCC=genu of corpus callosum; ICP=interior cerebellar peduncle; ML=medial lemniscus; MCP=middle cerebellar peduncle; PCT=pontine crossing tract; PCR=posterior corona radiata; PLIC=posterior limb of the internal capsule; PTR=posterior thalamic radiation; RLIC=retrolenticular part of the internal capsule; SCC=splenium of corpus callosum; SCR=superior corona radiata; SFO=superior fronto-occipital fasciculus; SLF=superior longitudinal fasciculus; SS=sagittal stratum; TAP=tapetum; UNC=uncinate fasciculus.

The HC group showed the most robust NSSD metrics in the left cuneus and inferior temporal gyrus, while the ROP group showed those in the left medial orbitofrontal, superior frontal, and fusiform gyri (Figure 2C). In the training sample (n’s as above), both groups showed a positive correlation between latent variables of GM thickness and WM FA with a greater correlation in controls (HC: *r*=0.43, *p*<0.001, ROP: *r*=0.32, *p*=0.007). The HC group only demonstrated a significant correlation in cross-validation data (n’s as above), and a generalisable correlation was not demonstrated in the out-of-sample data (n’s as above) (Figure 2D). GM and WM saliences showed convergence when sample size reached 60 per group (Figure 2E).

In summary, LV3 demonstrated a significant pattern derived from the relationship between GM thickness and WM FA and showing group differences. The pattern was strongly mapped onto ROP individuals, involving frontal and temporal GM regions and WM tracts, including the right corticospinal tract and bilateral anterior limb of the internal capsule. The salience patterns were stable in this sample size.

### MB-PLS-C between GM surface area and WM FA

The MB-PLS-C analyses between GM surface area and WM FA were significant (omnibus test *p*<0.0005). LV1 and LV2 were significant and explained 72.18% of the sum-of-squares variance (Figure S9 and Table S8).

### First latent variable (LV1)

LV1 explained 53.21% of sum-of-squares variance (*p*<0.0005), and both groups depicted a similar correlation pattern. The covariate blocks displayed a robust positive correlation between increased GM surface areas and increased total intracranial volume, which was also associated with male sex (Figure S9).

Significant GM saliences showed a pattern overlapping between the groups, with the controls demonstrating a stronger mapping than the ROP group (Figure 3A). In the HC group, the largest GM saliences were observed in temporal and parietal regions, whereas in the ROP group, those were observed in temporal and frontal regions. The largest positive WM saliences were observed in the right uncinate fasciculus, bilateral external capsule, and left superior corona radiata, and the largest negative saliences were found in the splenium of the corpus callosum and right anterior limb of the internal capsule (Figure 3B).

**Figure 3.**
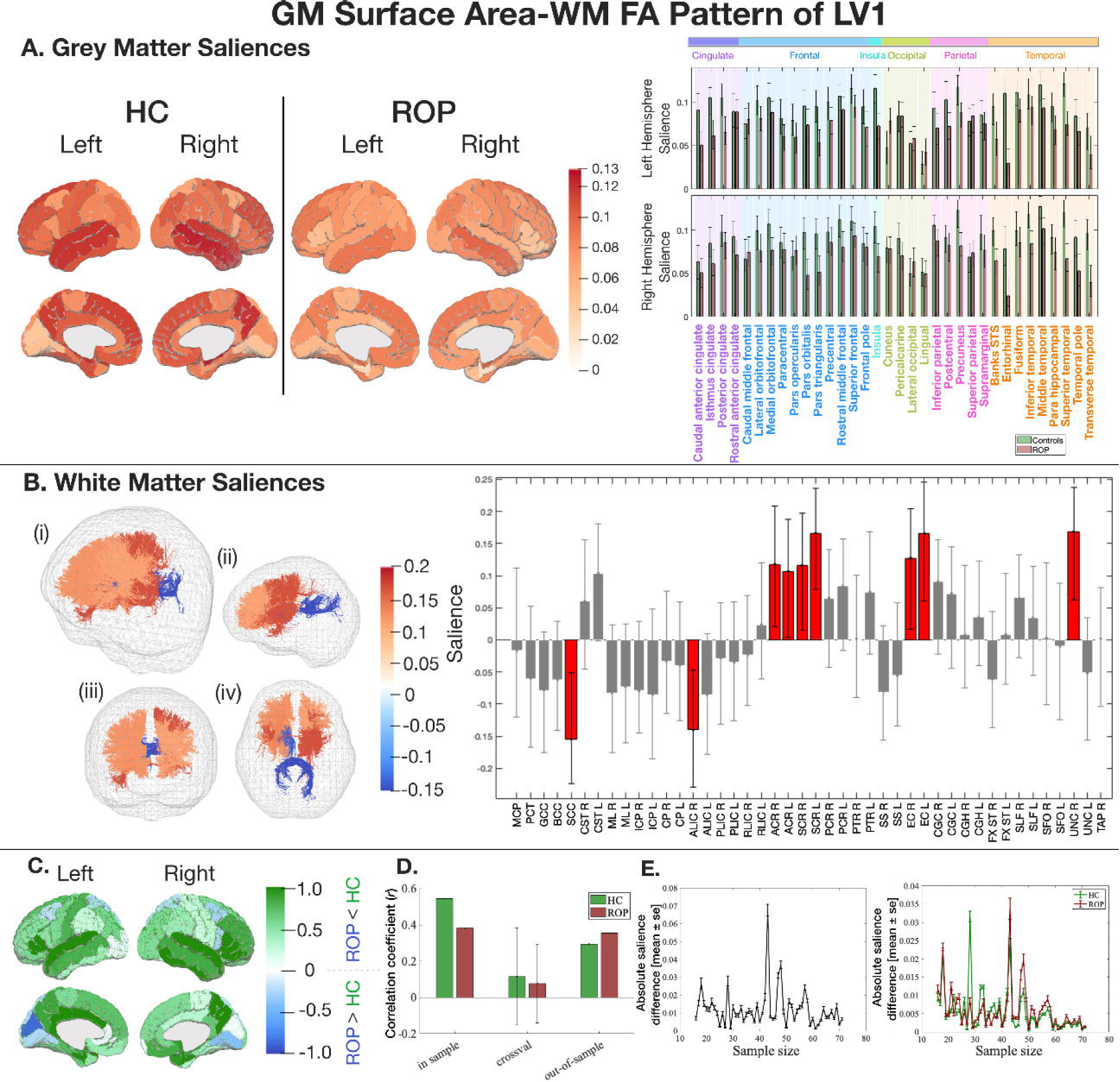
The latent pattern of LV1 between GM surface area and WM FA derived from MB-PLS-C. Key: A) GM salience by group in HC and ROP individuals. GM saliences of HC (green) and ROP (red) with 95% confidence interval (black lines) and color-coded lobe information. All the reliable saliences were positive. B) WM saliences shared between groups. WM tracts represented in tractography are colour-coded based on the salience intensity ((i) overall view (ii) from the left (iii) from the front (iv) from the bottom). The bar plot shows the salience of each WM tract with 95% confidence intervals (black lines). C) Normalised regional salience-differences. A positive score (green) shows a salience is larger in HC, and a negative score (blue) shows a salience is larger in ROP. D) A correlation coefficient between LV1 of GM and WM in the training and the test samples, and a correlation coefficient and its standard deviation from Monte Carlo cross-validation. Both groups displayed high correlations in the out-of-sample data, but neither group exhibited a significant correlation in the cross-validation data. E) The impact of sample size on salience intensity. Figures on the left and right depict differences in WM saliences and GM saliences, respectively. The GM-WM pattern converged with 50 samples from each group. ACR=anterior corona radiata; ALIC=anterior limb of the internal capsule; BCC=body of corpus callosum; CGC=cingulum (cingulate gyrus); CGH=cingulum (hippocampal portion); CP=cerebral peduncle; CR=corona radiata; CST=corticospinal tract; EC=external capsule; FX ST=fornix (cres) / stria terminalis; GCC=genu of corpus callosum; ICP=interior cerebellar peduncle; ML=medial lemniscus; MCP=middle cerebellar peduncle; PCT=pontine crossing tract; PCR=posterior corona radiata; PLIC=posterior limb of the internal capsule; PTR=posterior thalamic radiation; RLIC=retrolenticular part of the internal capsule; SCC=splenium of corpus callosum; SCR=superior corona radiata; SFO=superior fronto-occipital fasciculus; SLF=superior longitudinal fasciculus; SS=sagittal stratum; TAP=tapetum; UNC=uncinate fasciculus.

The HC group displayed the most robust NSSD metrics in the bilateral entorhinal cortex and right transverse temporal gyrus, while the ROP group showed those in the left cuneus and pericalcarine cortex and right lingual gyrus (Figure 3C). In the training sample (n’s as above), both groups showed a positive correlation between latent variables of GM surface area and WM FA, with larger correlations in the HCs (HC: *r*=0.54, *p*<0.001, ROP: *r*=0.38, *p*=0.001). Whilst both groups did not show significant correlations in cross-validation data (n’s as above), out-of-sample data (n’s as above) displayed similar correlations in both groups (Figure 3D). GM and WM saliences exhibited convergence when sample size reached 50 per group (Figure 3E).

In summary, LV1 represented a significant pattern derived from the relationship between GM surface area and intracranial volume and sex, overlapping between the ROP and HC groups. The pattern involved frontal, temporal, and parietal GM regions and WM tracts, including the right uncinate fasciculus and the splenium of the corpus callosum. The salience patterns were stable in this sample size.

### Second latent variable (LV2)

LV2 explained 18.97% of sum-of-squares variance (*p*=0.0003). The HC group demonstrated negative correlations between GM surface area and WM FA, while the ROP group exhibited positive correlations (Figure S9).

Significant GM saliences were in different directions between the groups, describing group differences (Figure 4A). The largest GM saliences were observed in cingulate, frontal, parietal, and temporal regions in controls, whereas those were observed in cingulate, frontal, and parietal regions in ROP individuals. All the WM saliences were positive with the largest saliences present bilaterally in the inferior cerebellar peduncle and posterior corona radiata and in the left superior corona radiata and superior longitudinal fasciculus (Figure 4B).

**Figure 4.**
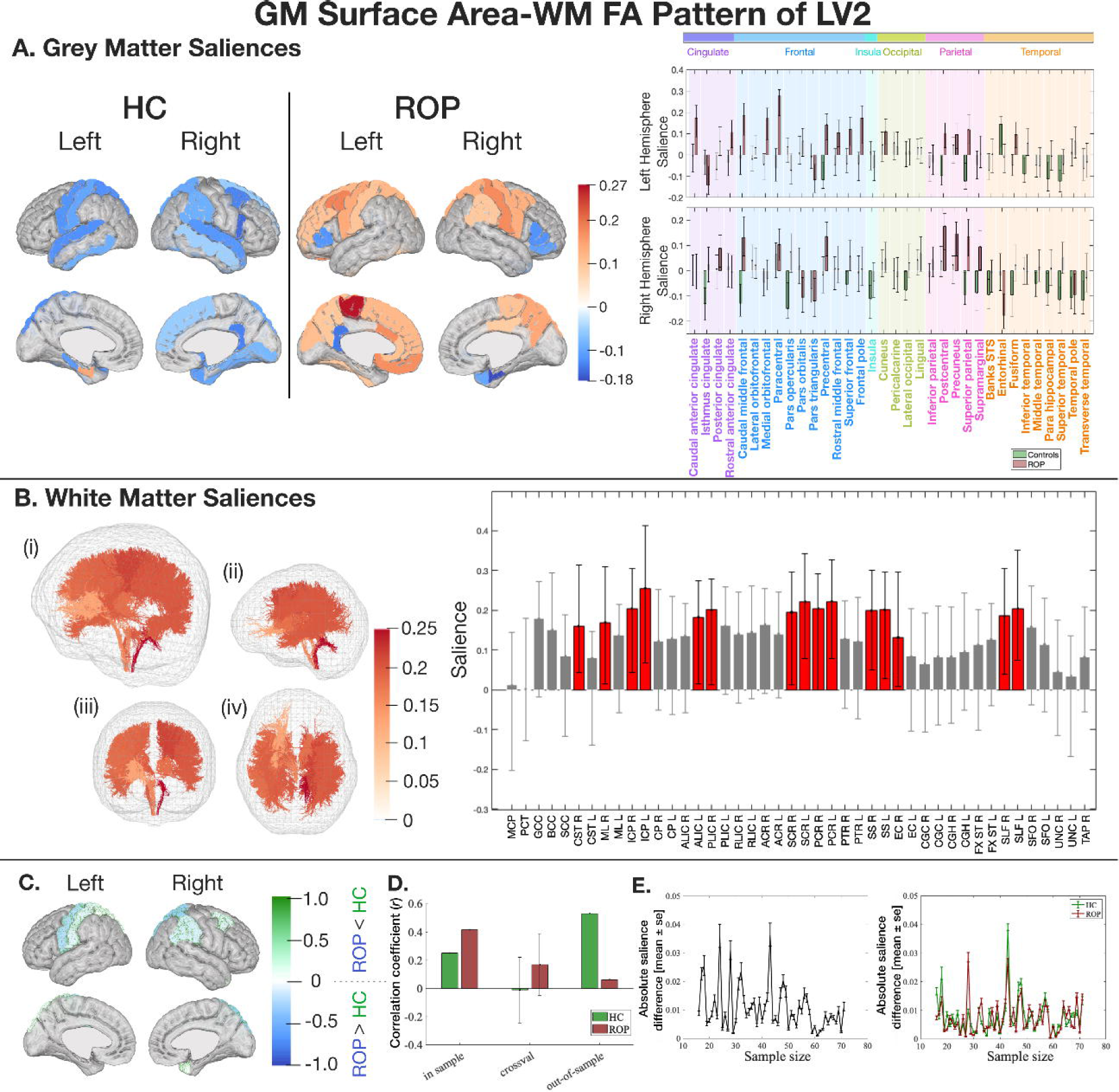
The latent pattern of LV2 between GM surface area and WM FA derived from MB-PLS-C. Key: A) GM salience by group in HC and ROP individuals. GM saliences of HC (green) and ROP (red) with 95% confidence interval (black lines) and color-coded lobe information. All the reliable saliences were positive. B) WM saliences shared between groups. WM tracts represented in tractography are colour-coded based on the salience intensity ((i) overall view (ii) from the left (iii) from the front (iv) from the bottom). The bar plot shows the salience of each WM tract with 95% confidence intervals (black lines). C) Normalised regional salience-differences. A positive score (green) shows a salience is larger in HC, and a negative score (blue) shows a salience is larger in ROP. D) A correlation coefficient between LV2 of GM and WM in the training and the test samples, and a correlation coefficient and its standard deviation from Monte Carlo cross-validation. Cross-validation data was unable to demonstrate a significant correlation, while out-of-sample data only showed a generalised correlation in the HC group and not in the ROP group. E) The impact of sample size on salience intensity. Figures on the left and right depict differences in WM saliences and GM saliences, respectively. The GM-WM pattern converged with 50 samples from each group. ACR=anterior corona radiata; ALIC=anterior limb of the internal capsule; BCC=body of corpus callosum; CGC=cingulum (cingulate gyrus); CGH=cingulum (hippocampal portion); CP=cerebral peduncle; CR=corona radiata; CST=corticospinal tract; EC=external capsule; FX ST=fornix (cres) / stria terminalis; GCC=genu of corpus callosum; ICP=interior cerebellar peduncle; ML=medial lemniscus; MCP=middle cerebellar peduncle; PCT=pontine crossing tract; PCR=posterior corona radiata; PLIC=posterior limb of the internal capsule; PTR=posterior thalamic radiation; RLIC=retrolenticular part of the internal capsule; SCC=splenium of corpus callosum; SCR=superior corona radiata; SFO=superior fronto-occipital fasciculus; SLF=superior longitudinal fasciculus; SS=sagittal stratum; TAP=tapetum; UNC=uncinate fasciculus.

The HC group showed the most robust NSSD metrics in the left superior parietal gyrus and right temporal pole, while the ROP group displayed those in the left precentral and right superior parietal and supramarginal gyri (Figure 4C). In the training sample (n’s as above), both groups showed positive correlations between each of the latent variables for GM surface area and WM FA, with a greater correlation in ROP (HC: *r*=0.25, *p=*0.033, ROP: *r*=0.42, *p*<0.001), but there were no significant correlations observed in the cross-validation data (n’s as above). Out-of-sample data (n’s as above) demonstrated a generalised correlation only in the HC group but not in the ROP group (Figure 4D). GM and WM saliences converged after 50 samples per group (Figure 4E).

In summary, LV2 showed a significant pattern, derived from the relationship between GM surface area and WM FA, that shows group differences. The pattern involved cingulate, frontal, parietal, and temporal GM regions and WM tracts, including the bilateral inferior cerebellar peduncle and posterior corona radiata. The salience patterns were stable in this sample size.

### Tractography-based examination of GM-WM patterns derived from MB-PLS-C

To assess the anatomical plausibility of GM-WM patterns showing group differences, we calculated the strength of direct cortico-cortical connectivity within the GM-WM patterns using tractography and performed a permutation test to evaluate whether the strength of direct cortico-cortical connectivity was significantly high. The test demonstrated significantly high direct cortico-cortical connectivity in the ‘GM surface area’-‘WM FA’ pattern (*p*=0.0002 (ROP), *p*=0.012 (HC)) but not in the ‘GM thickness’-‘WM FA’ pattern (*p*=0.87 (ROP), *p*=0.72 (HC)). Figure 5 visualises the direct cortico-cortical connections in the GM-WM patterns showing group differences. As demonstrated by permutation testing, the ‘GM thickness’-‘WM FA’ pattern was based on relatively low direct cortico-cortical connectivity, while the ‘GM surface area’-‘WM FA’ pattern was based on high direct cortico-cortical connectivity between GM regions. Figure 5B shows that the ‘GM thickness’-‘WM FA’ pattern has relatively high cortico-subcortical connectivity compared with cortico-cortical connectivity.

**Figure 5.**
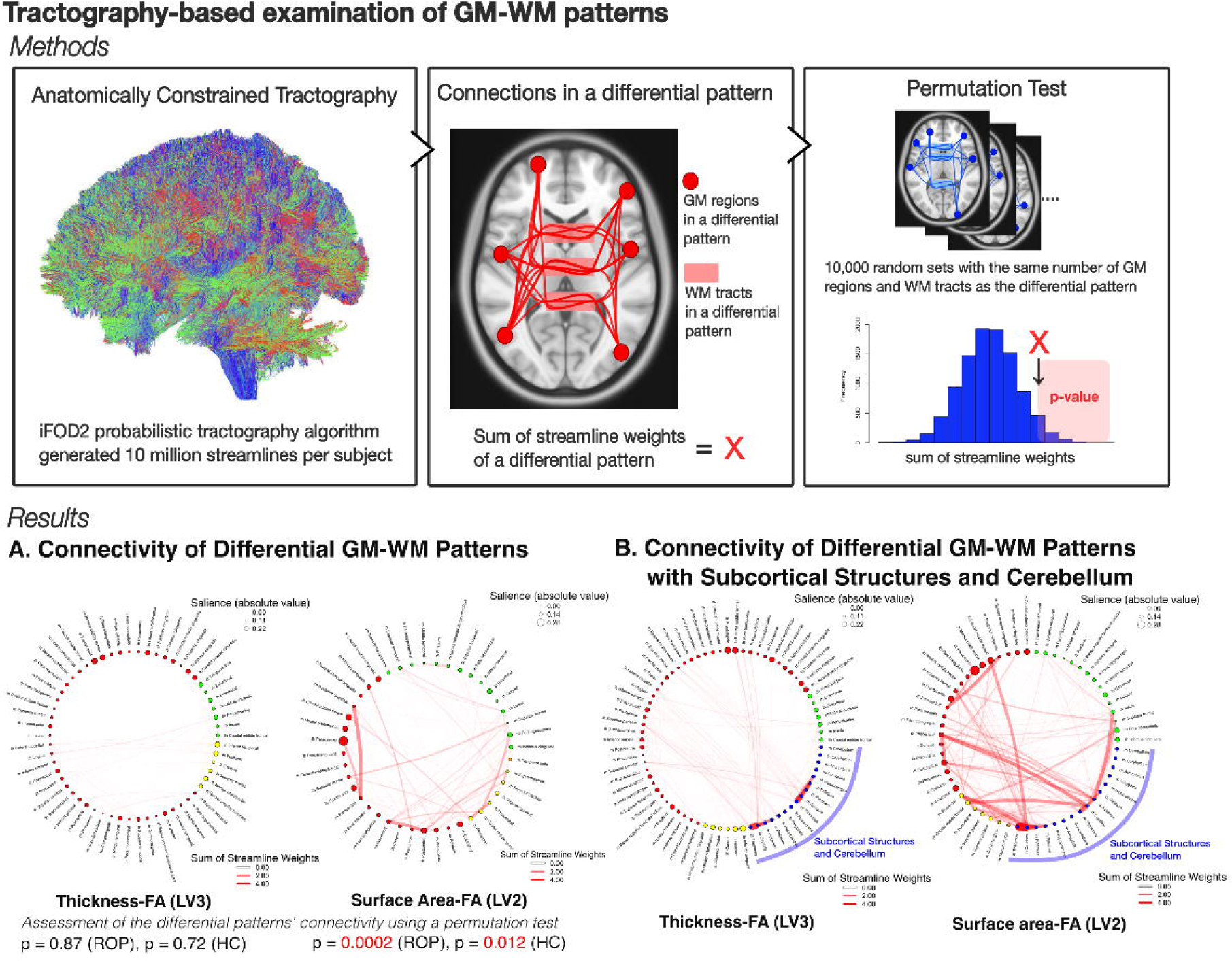
The tractography-based connectivity of GM and WM with a significant salience. Key: Connectograms in the result section show the connectivity of differential patterns with a significant salience. Circles represent GM regions (red: regions with a significant salience in ROP, green: regions with a significant salience in controls, yellow: regions with saliences of different directions between groups, orange: regions with saliences of the same direction between groups, blue: subcortical regions and cerebellum). The size of a circle exhibits the salience size of a GM region (subcortical regions and cerebellum are assigned a fixed value), and edges represent the sum of streamline weights between GM regions. Result A) Connectograms illustrate the connectivity of differential patterns. Compared to the ‘GM surface area’-‘WM FA’ pattern, the ‘GM thickness’-‘WM FA’ pattern consists of connections with a smaller sum of streamline weights. Result B) Connectograms display the connectivity of differential patterns with subcortical structures and cerebellum. In the ‘GM thickness’-‘WM FA’ pattern, cortical regions with a significant salience have relatively high connectivity with subcortical regions.

### Correlations between cognitive abilities and latent variables of GM-WM patterns describing group differences

In the ‘GM thickness’-‘WM FA’ pattern demonstrating group differences, with GM saliences in different directions (LV3), the ROP group showed a significant correlation between the WM latent variables and processing speed (ρ=0.32, *p*=0.006, *p_adjusted_*=0.048). The HC group did not exhibit a significant correlation. The Fisher’s z-test further demonstrated a significant group difference in a correlation between the WM latent variables and processing speed (z=2.34, *p*=0.019). The details are provided in the Supplementary Material.

In the ‘GM surface area’-‘WM FA’ pattern representing group differences (LV2), only word reading ability showed a significant correlation with the GM latent variables in the ROP group, which did not survive multiple comparison corrections (ρ=0.24, p=0.047, *p_adjusted_*=0.30).

### Correlations between the latent variables and PANSS scores and doses of antipsychotics

PANSS negative scores were correlated with the WM latent variables of the ‘GM thickness’-‘WM FA’ pattern showing group differences (LV3) (ρ*=*0.251*, p*=0.038, *p_adjusted_*=0.23) and the WM latent variables of the ‘GM surface area’-‘WM FA’ pattern describing group differences (LV2) (ρ=-0.245, *p*=0.042, *p_adjusted_*=0.25) in ROP. However, none of these correlations was significant after the Benjamini-Hochberg method with a 5% false discovery rate ([3 PANSS scores] × [GM or WM]). The latent variables derived from MB-PLS-C were not significantly correlated with doses of antipsychotics.

### Comparisons between latent salience patterns for GM thickness and surface area derived from MB-PLS-C and allometric scaling maps

Allometric scaling maps were employed to investigate the similarity between latent cortical patterns identified through MB-PLS-C analysis and naturally occurring cortical variations [63]. ‘GM thickness’-‘WM FA’ salience patterns demonstrated significant correlations with the allometric scaling maps in both groups in LV2 and 3. ‘GM surface area’-‘WM FA’ salience patterns also exhibited significant correlations with the allometric scaling maps, except in the left hemisphere pattern of the ROP group in LV2. Details are described in the Supplementary Material.

## DISCUSSION

This is the first study to examine 1) multivariate grey matter-white matter (GM-WM) patterns in recent-onset psychosis (ROP) individuals and healthy controls (HC) using multiblock partial least squares correlation (MB-PLS-C) and 2) associations between these patterns and cognitive abilities. MB-PLS-C identified a ‘GM thickness’-‘WM fractional anisotropy (FA)’ pattern strongly mapped onto the HC group, a ‘GM surface area’-‘WM FA’ pattern overlapping between groups, and ‘GM surface area’-‘WM FA’ and ‘GM thickness’-‘WM FA’ patterns showing group differences. The ‘GM thickness’-‘WM FA’ pattern representing group differences was associated with frontal and temporal GM regions in the ROP group, including the left medial orbitofrontal, superior frontal, fusiform, and inferior temporal gyri, and WM tracts, including the corticospinal tract and anterior limb of the internal capsule. The ‘GM surface area’-‘WM FA’ pattern demonstrating group differences was associated with cingulate, frontal, temporal, and parietal GM regions and WM tracts, including the inferior cerebellar peduncle, superior and posterior corona radiata, and superior longitudinal fasciculus. Processing speed was significantly correlated with the ‘GM thickness’-‘WM FA’ pattern which showed group differences. These findings suggest potential signatures of aberrant ‘GM thickness’-‘WM FA’ and ‘GM surface area’-‘WM FA’ changes in the early stages of schizophrenia-spectrum disorders, with an association with cognitive abilities.

### ‘GM thickness’-‘WM FA’ patterns

The pattern (2nd latent variable) explaining 16.9% of the variance in the ‘GM thickness’-‘WM FA’ analysis identified negative thickness-age correlations only in controls. This pattern, involving the dorsolateral and medial prefrontal cortex, is consistent with previous longitudinal studies indicating that typical age-related reductions in cortical thickness progress from more posterior cortical areas to more anterior associative cortical regions later in adolescence [64–66]. ROP individuals did not demonstrate a similar pattern, suggesting that the disorder may disrupt the relationship between ‘GM thickness’ and age observed in controls.

The ‘GM thickness’-‘WM FA’ pattern representing group differences (3rd latent variable) explained 12.4% of the variance, was strongly associated with ROP individuals and largely involved frontal and temporal GM regions, which are consistently implicated in schizophrenia [1,4,5]. This pattern was associated with robust ‘WM FA’ contributions in seven tracts, with the highest contributions in the right corticospinal tract as well as the anterior limb and retrolenticular part of the internal capsule. This GM-WM pattern indicates that these WM tracts, which connect the cortex with subcortical structures and the spinal cord, are associated with frontal and temporal GM changes in schizophrenia-spectrum disorders. To our knowledge, no studies have examined multivariate relationships of WM tracts with cortical thickness. Our method using MB-PLS-C identified a WM pattern not previously identified in studies examining WM structural differences in early-stage schizophrenia [67,68]. Considering that GM thickness is influenced by environment and undergoes continual changes through adulthood, this pattern may reflect changes in WM that are linked with changes in GM thickness occurring around the onset of psychosis [69–71].

### ‘GM surface area’-‘WM FA’ patterns

The shared ‘GM surface area’-‘WM FA’ pattern (1st latent variable, explaining 53.2%) identified by MB-PLS-C was mainly driven by positive correlations between GM surface area and its covariates, namely, intracranial volume and sex. This is indicative of general associations between a larger brain surface area and a larger intracranial volume and male sex in both groups. Examination of surface area is relevant to understanding GM-WM relationships during early neurodevelopment as surface area is governed by the migration of neurons early in life [69,70,72]. The smaller contributions in the GM pattern in the ROP group compared to controls suggest a reduced association between surface area and its covariates in ROP, indicating a normal but attenuated pattern of neurodevelopment in ROP, consistent with the neurodevelopmental hypothesis of schizophrenia [73,74].

The ‘GM surface area’-‘WM FA’ pattern exhibiting group differences (2nd latent variable, explaining 19.0%) involved positive contributions in the ROP group mainly for left frontal and bilateral parietal regions and negative contributions in controls mainly for right frontal and temporal and bilateral parietal regions. Considering additional analyses demonstrated that the GM pattern in the left hemisphere of the ROP group was not correlated with allometric scaling maps, these findings imply the absence of neurodevelopment-associated GM-WM correlations in schizophrenia-spectrum disorders. WM tracts involved in this pattern include the inferior cerebellar peduncle, superior and posterior corona radiata, superior longitudinal fasciculus, and sagittal stratum. The superior longitudinal fasciculus and sagittal stratum reach maturity earlier in patients than in controls, as demonstrated by Cetin-Karayumak et al. (2020), suggesting a relationship between the GM-WM pattern and neurodevelopment [75]. Interestingly, the inferior cerebellar peduncle, conveying proprioceptive sensory input to the cerebellum, showed the largest contribution to this pattern reflecting group differences. While few studies have investigated this WM tract in schizophrenia-spectrum disorders, changes in the inferior cerebellar peduncle correlated with GM regions may be associated with sensory prediction deficits in schizophrenia-spectrum disorder [76–78].

### Identification of a ‘GM thickness’-‘WM FA’ pattern independent of direct connections by MB-PLS-C

We examined the anatomical plausibility of the GM-WM patterns showing group differences by calculating network connectivity within these patterns using WM tractography and also compared the connectivity with and without subcortical regions included. The results suggest that ‘GM thickness’-‘WM FA’ pattern was not reflected by direct cortico-cortical connections but rather by indirect pathways, involving subcortical structures, including thalamus, striatum, and cerebellum, consistent with functional imaging studies [79–82]. These findings highlight the efficacy of MB-PLS-C in exploring indirectly correlated GM-WM relationships, which are not readily observed with conventional approaches. MB-PLS-C’s potential is demonstrated in mapping multivariate whole-brain GM-WM ‘signatures’ at the earliest stages of psychosis.

### Association between GM-WM relationships and cognitive abilities

In ROP individuals, the WM component of 3rd latent variable (the ‘GM thickness’-‘WM FA’ pattern exhibiting group differences) was significantly correlated with processing speed. This correlation is consistent with past studies showing a strong association between processing speed and WM microstructure changes in schizophrenia-spectrum disorders [83–87]. Processing speed is a core cognitive deficit in schizophrenia-spectrum disorders and has been shown to mediate the relationship between WM changes and other cognitive abilities, such as working memory, in these disorders [50,84,86,88]. This relationship of processing speed to GM thickness, rather than GM surface area, and the associated WM FA are relevant to the early stages of psychosis as GM thickness continues to change from adolescence to early adulthood in healthy individuals with evidence for accelerated change in the early stages of schizophrenia [89,90]. Further, WM changes appear to follow GM changes developmentally as schizophrenia develops into a chronic illness [13,91]. In contrast, surface area is determined in early neurodevelopment [69,71]. This suggests the alterations in WM FA in relation to GM thickness changes around the onset may be driving the observed low processing speed. However, longitudinal studies are needed to assess such a causal relationship.

### Further analysis, limitations, and conclusion

PANSS negative scores were associated with the GM thickness’-‘WM FA’ and ‘GM surface area’-‘WM FA’ patterns representing group differences. The association between PANSS negative scores and brain alterations has been demonstrated in a previous PLS study and a recent review paper [22,92]. The present study demonstrated the close association between negative symptoms and GM-WM changes in schizophrenia. However, it is important to note that these correlations did not survive multiple comparison corrections.

Our study is subject to several limitations. Firstly, the antipsychotic medication effect on GM and WM changes was not accounted for. While no significant correlations between latent variables and medication doses were observed, further study with antipsychotic-naive individuals is needed. Secondly, other confounders such as substance use, illness duration, and comorbid mental and physical illnesses were not considered, despite adjusting for gender, age, intracranial volume, and head motion within the scanner in the current MB-PLS-C. This study employed the MB-PLS-C analysis with a large number of variables, leveraging PLS-C’s ability to select only significant components in the model, which is consistent with practices in previous research. While the analysis using cross-validation and out-of-sample did not prove the generalisability of the models, the results may reflect the heterogeneity in a schizophrenia-spectrum disorders population, especially in the early stages of illness [93–96]. The differences in duration of onset and country between the out-of-sample analysis and the main sample may also explain the failure to reproduce the main sample’s patterns. Further GM-WM MB-PLS-C studies with a larger sample size or in a different population would provide insights into the heterogeneity of brain patterns in individuals with early psychosis.

In summary, our MB-PLS-C approach identified correlated whole-brain patterns of ‘GM thickness’-‘WM FA’ and ‘GM surface area’-‘WM FA’ in ROP individuals compared to controls. The ‘GM thickness’-‘WM FA’ pattern showing group differences observed in the MB-PLS-C exhibited the most pronounced contribution in frontal and temporal lobes and WM tracts, such as the corticospinal tract and anterior limb of the internal capsule. For the ‘GM surface area’-‘WM FA’ pattern representing group differences, the most significant contribution was observed in frontal, temporal, and parietal regions and WM tracts, such as the inferior cerebellar peduncle and superior corona radiata. The ‘GM thickness’-‘WM FA’ pattern describing group differences demonstrated a significant association with processing speed. Our findings of a relationship between cognitive function and GM-WM patterns for GM thickness rather than for surface area have implications for our understanding of brain-behaviour relationships neurodevelopmentally in psychosis. Further studies on ultra-high-risk and chronic schizophrenia individuals are required to track patterns showing group differences and their progression over the course of illness. Identifying these patterns and their association with cognitive abilities in the early stages around the onset would provide information relevant to early interventions for cognitive impairments of schizophrenia-spectrum disorders.

## DATA AVAILABILITY

Brain imaging and cognitive and clinical data from the Human Connectome Project for early psychosis can be requested from https://www.humanconnectome.org/study/human-connectome-project-for-early-psychosis. Brain imaging and cognitive data from the Human Connectome Project Development can be requested from https://www.humanconnectome.org/study/hcp-lifespan-development. Access to these datasets requires approval. Other data can be obtained from the corresponding author upon reasonable request.

## CODE AVAILABILITY

MB-PLS-C was performed in Matlab 2022b. Codes can be available from the corresponding author upon reasonable request.

## ACKNOWLEDGEMENTS

Research using Human Connectome Project for Early Psychosis (HCP-EP) data reported in this publication was supported by the National Institute of Mental Health of the National Institutes of Health under Award Number U01MH109977. The HCP-EP 1.1 Release data used in this report came from DOI: 10.15154/1522899. YS was supported by the Research Training Program Scholarship, Heiwa Nakajima Foundation scholarship, and Japan Student Services Organization Scholarship. CP was supported by a National Health and Medical Research Council (NHMRC) Investigator Grant (ID: 1196508) and NHMRC Program Grant (ID: 1150083). VC was supported by an NHMRC Investigator Grant (ID: 1177370) and University of Melbourne Dame Kate Campbell Fellowship.

## CONTRIBUTION

YS contributed to planning experiments, developed the MB-PLS-C code, conducted image processing and all analyses, and prepared, reviewed and edited the manuscript. CP proposed the study, contributed to planning experiments and reviewed and edited the manuscript. VC contributed to the collection and processing of out-of-sample data and reviewed and edited the manuscript. LL contributed to the collection and processing of out-of-sample data and reviewed the manuscript. CMW conceptualised the study, contributed to planning experiments, and reviewed and edited the manuscript. WTS conceptualised the study, designed the MB-PLS-C code, provided a technical support, contributed to planning experiments, and reviewed and edited the manuscript.

Supplementary information is available at MP’s website.

