## Supplementary Materials for "Characterising grey-white matter relationships in recent-onset psychosis and its association with cognitive function"

***Supplementary Material***

**MATERIALS AND METHODS	3**

Participants	3

Figure S1. Flowchart of subject selection.	4

Table S1. Demographic and clinical data of subjects before propensity score matching.	5

Table S2. NIH Toolbox Cognitive Battery.	6

Clinical Assessment	6

MRI acquisition	6

MRI processing	6

Tract-Based Spatial Statistics	7

Figure S2. The sum of squared errors in diffusion tensor estimation.	8

Harmonisation	8

Table S3. Site information.	8

Figure S3. Harmonisation on GM variables.	10

Figure S4. Harmonisation on WM variables.	11

Table S4. Age and sex of controls recruited for harmonisation with RISH features	11

Tractography	12

Statistical analyses	12

Multiblock partial least squares correlation analyses (MB-PLS-C)	12

Figure S5: The input cross-correlation matrix for MB-PLS-C analysis.	13

Table S5. Subject information of out-of-sample	15

Normalised between-group structural salience difference (NSSD)	15

Sample size and salience convergence	16

Comparison between latent cortical patterns and allometric scaling maps	16

**RESULTS	17**

Case-control differences in GM and WM	17

Figure S6. Group differences in GM variables between recent onset psychosis and healthy controls.	17

Figure S7. Group differences in WM variables between recent onset psychosis and healthy controls.	19

Table S6. Group differences in cognitive variables.	20

PLS between GM thickness and WM FA	20

Figure S8. Decomposition of the input multi-block cross-correlation matrix by MB-PLS-C between GM thickness and WM FA.	20

Table S7. MB-PLS-C latent variables contributing to the relationship between GM thickness and WM FA.	20

PLS between GM surface area and WM FA	22

Figure S9. Decomposition of the input multi-block cross-correlation by MB-PLS-C between GM surface area and WM FA.	22

Table S8. MB-PLS-C latent variables contributing to the relationship between GM surface area and WM FA.	22

PLS between GM volume and WM FA	23

Figure S10. Decomposition of the input multi-block cross-correlation by MB-PLS-C between GM volume and WM FA.	23

Table S9. MB-PLS-C latent variables contributing to the relationship between GM volume and WM FA.	23

Figure S11. The latent pattern of LV1 between GM volume and WM FA derived from MB-PLS-C.	25

Figure S12. The latent pattern of LV2 between GM volume and WM FA derived from MB-PLS-C.	28

PLS between GM volume and WM MD	30

Figure S13. Decomposition of the input multi-block cross-correlation matrix by MB-PLS-C between GM volume and WM MD.	30

Table S10. MB-PLS-C latent variables contributing to the relationship between GM volume and WM MD.	30

Figure S14. The latent pattern of LV1 between GM volume and WM MD derived from MB-PLS-C.	32

Figure S15. The latent pattern of LV2 between GM volume and WM MD derived from MB-PLS-C.	35

PLS between GM thickness and WM MD	37

Figure S16. Decomposition of the input multi-block cross-correlation matrix in by MB-PLS-C between GM thickness and WM MD.	37

Table S11. MB-PLS-C latent variables contributing to the relationship between GM thickness and WM MD.	37

Figure S17. The latent pattern of LV3 between GM thickness and WM MD derived from MB-PLS-C.	39

PLS between GM surface area and WM MD	41

Figure S18. Decomposition of the input multi-block cross-correlation matrix by MB-PLS-C between GM surface area and WM MD.	41

Table S12. MB-PLS-C latent variables contributing to the relationship between GM surface area and WM MD.	41

Figure S19. The latent pattern of LV1 between GM surface area and WM MD derived from MB-PLS-C.	43

Two-group input multiblock cross-correlation Matrix, R	45

Figure S20. The multiblock cross-correlation matrix presenting the correlations between GM volume, WM FA, and their covariate blocks.	45

Figure S21. The multiblock cross-correlation matrix presenting the correlations between GM thickness, WM FA, and their covariate blocks.	46

Figure S22. The multiblock cross-correlation matrix presenting the correlations between GM surface area, WM FA, and their covariate blocks.	47

Figure S23. The multiblock cross-correlation matrix presenting the correlations between GM volume, WM MD, and their covariate blocks.	48

Figure S24. The multiblock cross-correlation matrix presenting the correlations between GM thickness, WM MD, and their covariate blocks.	49

Figure S25. The multiblock cross-correlation matrix presenting the correlations between GM surface area, WM MD, and their covariate blocks.	50

Correlations between cognitive abilities and latent variables of GM-WM patterns describing group differences	51

Correlations between allometric scaling maps and latent GM thickness and GM surface area patterns	51

### MATERIALS AND METHODS

#### Participants

The study consisted of recent-onset psychosis (ROP) individuals from the Human Connectome Project for Early Psychosis (HCP-EP) and healthy controls (HCs) from two datasets, HCP-EP and the Human Connectome Project in Development (HCP-D) [[1, 2]](https://paperpile.com/c/iqLHPy/li7H1+GpIQG). HCP-EP collected clinical and cognitive data and T1-weighted and diffusion MRI scans from 126 ROP individuals diagnosed with a schizophrenia spectrum disorder according to the DSM-5, within five years of their first psychotic episode, along with 68 matched controls. HCP-D collected data from HCs with the same cognitive data and T1-weighted and diffusion MRI scans as HCP-EP. We included 214 subjects from Harvard University out of the four imaging sites of HCP-D to minimise non-biological variability caused by imaging site differences.

Due to differences in the inclusion and exclusion criteria for HCs between the HCP datasets, we included those from HCP-D who did not take psychiatric medication and did not have parents diagnosed with schizophrenia spectrum disorders. Considering the typical period of psychosis onset, we included subjects between the ages of 16 and 30 [[3]](https://paperpile.com/c/iqLHPy/3YKVZ). The subject selection process is depicted in Figure S1, and their demographic and clinical details are presented in Table S1. To match the number of subjects, sex, and age between groups from the two datasets, propensity score matching was performed on pooled HCs [[4, 5]](https://paperpile.com/c/iqLHPy/wymDa+aghUe). We calculated the propensity score of each subject using a logistic regression model with sex and age as covariates and performed a nearest-neighbour matching algorithm without replacement based on the propensity score. We confirmed the matching achieved adequate balance by verifying that the standardised mean differences of sex and age were below 0.1 (Table 1).

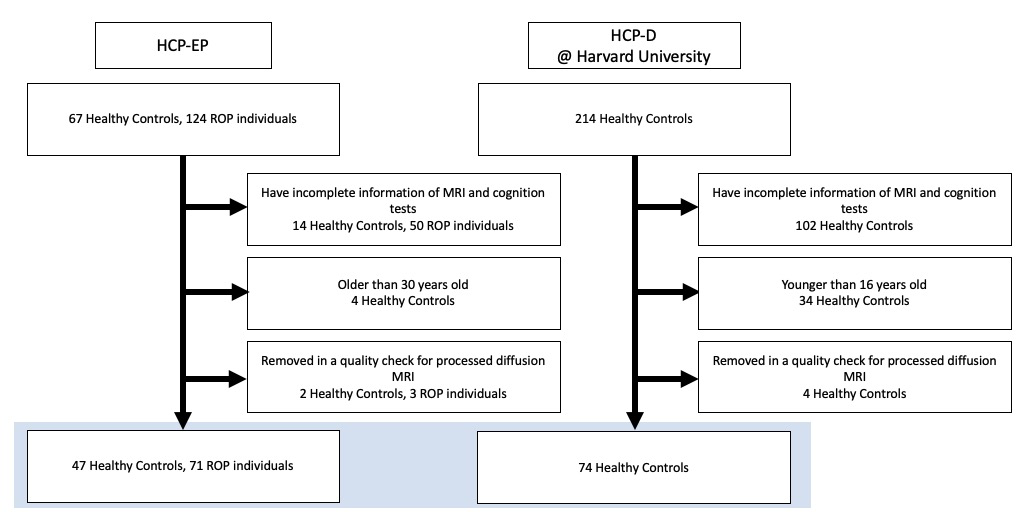

##### Figure S1. Flowchart of subject selection.

Key: The study selected subjects who provided complete information of MRI scans and cognitive abilities and were aged between 16 and 30 years.

##### Table S1. Demographic and clinical data of subjects before propensity score matching.

|  |  | Human Connectome Project for Early Psychosis (HCP-EP) | | | | |  | Human Connectome Project in Development  (HCP-D) | |
| --- | --- | --- | --- | --- | --- | --- | --- | --- | --- |
| Variables |  | Healthy Controls (*n* = 49) | |  | Non-affective psychotic individuals (*n* = 71) | |  | Healthy Controls (*n* = 74) | |
|  |  | **n** | **%** |  | **n** | **%** |  | **n** | **%** |
| Sex, male |  | 30 | 63.83 |  | 49 | 69.01 |  | 34 | 45.95 |
|  |  | **Mean** | **SD** |  | **Mean** | **SD** |  | **Mean** | **SD** |
| Age |  | 24.3 | 3.1 |  | 22.1 | 3.1 |  | 18.9 | 1.7 |
| PANSS* |  |  |  |  | **Mean** | **SD** |  |  |  |
| Positive |  |  |  |  | 11.3 | 4.2 |  |  |  |
| Negative |  |  |  |  | 15.3 | 5.9 |  |  |  |
| General |  |  |  |  | 25.4 | 6.7 |  |  |  |
| Total |  |  |  |  | 52.0 | 13.2 |  |  |  |
| Primary Lifetime Diagnosis |  |  |  |  | **n** | **%** |  |  |  |
| Schizophrenia |  |  |  |  | 51 | 71.83 |  |  |  |
| Schizoaffective Disorder |  |  |  |  | 9 | 12.68 |  |  |  |
| Schizophreniform Disorder |  |  |  |  | 8 | 11.27 |  |  |  |
| Other non-affective psychotic disease |  |  |  |  | 3 | 4.23 |  |  |  |
| Bipolar I Disorder |  |  |  |  |  |  |  |  |  |
| Major Depressive Disorder |  |  |  |  |  |  |  |  |  |
| GAF |  | **Mean** | **SD** |  | **Mean** | **SD** |  |  |  |
| Symptomatic functioning |  | 91.28 | 8.77 |  | 60.62 | 17.86 |  |  |  |
| Social functioning |  | 92.02 | 6.62 |  | 62.59 | 23.86 |  |  |  |
| Occupational functioning |  | 89.06 | 12.03 |  | 68.55 | 14.78 |  |  |  |

Key: *Two ROP individuals were excluded because of incomplete data.

##### Table S2. NIH Toolbox Cognitive Battery.

| **Cognitive tests** | **Variables used** | **Cognitive functions assessed** |
| --- | --- | --- |
| List Sorting Working Memory Test | Total number of items correctly recalled (0-26) | Working memory |
| Picture Sequence Memory Test | Theta score* | Episodic memory |
| Pattern Comparison Processing Speed Test | Total number of items correctly answered (0-130) | Processing speed |
| Oral Reading Recognition Test | Theta score* | Word reading skill |

Key: *Theta score is a score based on Item Response Theory (IRT) similar to the z-score, representing the relative overall ability or performance of the participant.

#### Clinical Assessment

The measures of clinical symptoms for the ROP group of HCP-EP included the Positive and Negative Syndrome Scale (PANSS) and the MIRECC Global Assessment of Functioning (GAF) [[6, 7]](https://paperpile.com/c/iqLHPy/Wlo0d+78IS9).

#### MRI acquisition

The subject data were downloaded from the HCP consortium. T1-weighted and diffusion images of HCP-EP were collected with 3T Siemens MAGNETOM Prisma scanners at Indiana University, Brigham and Women's Hospital, and McLean Hospital, and those of HCP-D were with 3T Siemens MAGNETOM Prisma scanners at Harvard University. Sequence parameters for both datasets were similar:

[T1-weighted images] 0.8 mm isotropic voxels, field of view = 256 x 256 mm^2^, repetition time = 2400 ms, echo time = 2.22 ms, and flip angle = 8° (HCP-EP):0.8 mm isotropic voxels, field of view = 256 x 256 mm^2^, repetition time = 2500 ms, echo time = 1.8/3.6/5.4/7.2 ms, and flip angle = 8° (HCP-D).

[Diffusion images] 1.5 mm isotropic voxels, field of view = 210 x 210 mm^2^, repetition time = 3230 ms, echo time = 89.20 ms, and flip angle = 78°, axial slice orientation with 92 slices and no gaps, 99 isotropically distributed diffusion-weighted directions in each shell (b=1500 and 3000 seconds/mm2) (HCP-EP/HCP-D).

#### MRI processing

Preprocessing on T1-weighted and diffusion-weighted MRI data was performed with the minimal preprocessing pipeline [[8]](https://paperpile.com/c/iqLHPy/22QLL). The HCP consortium preprocessed T1-weighted images, and the pipeline included the correction of MR gradient-nonlinearity-induced distortions, motion correction, intensity normalisation, skull strip, and cortical parcellation using the Desikan-Killiany (DK) Atlas with FreeSurfer (v6.0.0) ([https://surfer.nmr.mgh.harvard.edu](https://surfer.nmr.mgh.harvard.edu/)). GM volume (68 cortical regions, 14 subcortical structures, and the cerebellum cortex), thickness, and surface area (68 cortical regions) were computed [[9]](https://paperpile.com/c/iqLHPy/0eMQT).

We preprocessed the diffusion-weighted images with the pipeline, including EPI distortion, eddy current, and motion correction with FSL (FMRIB's Software Library, www.fmrib.ox.ac.uk/fsl) (v6.0.4). The quality of preprocessed images was visually inspected with two quality check frameworks of FSL (v6.0.4), QUAD (Quality Assessment for DMRI) and SQUAD (Study-wise Quality Assessment for DMRI) for diffusion-weighted images. Seven subjects from HCP-EP and four from HCP-D were excluded from the analysis due to the inadequate preprocessing quality [[10]](https://paperpile.com/c/iqLHPy/J9rhS).

#### Tract-Based Spatial Statistics

A tract-based spatial statistics (TBSS) analysis was conducted to determine WM parameters, fractional anisotropy (FA) and mean diffusivity (MD), as described by Smith et al. (2006) [[11]](https://paperpile.com/c/iqLHPy/5mIlo). WM value maps were generated using DTIFIT, and the FA maps of each subject were nonlinearly registered to the MNI152 template with a resolution of 1x1x1mm^3^. A mean FA map was produced from all the images, and a skeleton was formed with a threshold at an FA value of 0.2. The FA values for each white matter pathway were computed using the skeleton and the Johns Hopkins University (JHU) white matter tract atlas. For non-FA metrics, the "tbss_non_FA" command in FSL nonlinearly registered the maps to the template, and the WM values were calculated for each subject's white matter pathways. WM information, including FA and MD, was computed for 48 regions of the JHU white matter tract atlas [[12]](https://paperpile.com/c/iqLHPy/fjHQC).

The fornix, left tapetum, and superior cerebellar peduncles were removed from the investigation due to the large sum of squared errors in diffusion tensor estimation (Figure S2).

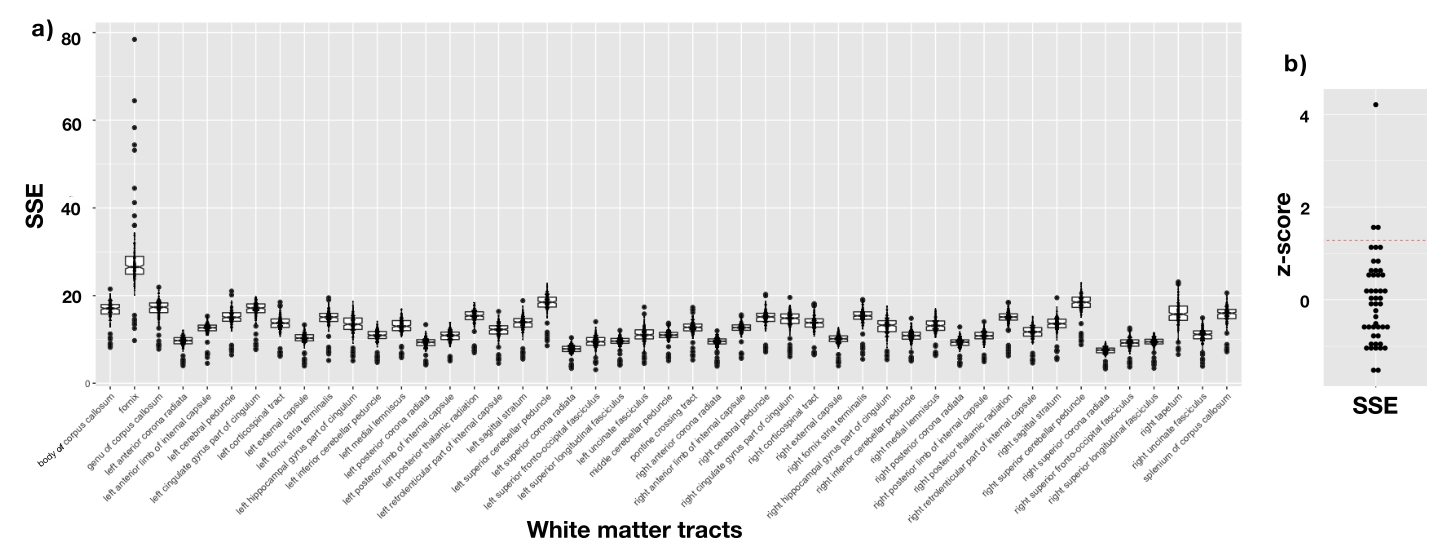

##### Figure S2. The sum of squared errors in diffusion tensor estimation.

Key: a) The average tract-wise sum of squared error (SSE) for tensor estimation was computed using FSL. The box plot illustrates each tract’s median, quartiles, and outliers, and the dot plot depicts each subject. b) The average tract-wise SSE of all subjects was transformed into a z-score with a threshold at 1.281 (red line, significance level of 10% in a one-tailed test). The left tapetum exhibited a noticeably large deviation and was excluded before plotting. The fornix and superior cerebellar peduncles on both sides exceeded the threshold and were excluded from the present analysis.

#### Harmonisation

The neuroimaging data were obtained from four different imaging sites, requiring GM and WM variables to be harmonised to reduce the effect of the scanner and protocol differences (Table S3).

##### Table S3. Site information.

| Variables |  | Healthy Controls (*n* = 71) | |  | ROP individuals (*n* = 71) | |
| --- | --- | --- | --- | --- | --- | --- |
| Site |  | **n** | **%** |  | **n** | **%** |
| Indiana University |  | 15 | 21.13 |  | 45 | 63.38 |
| Brigham and Women's Hospital |  | 14 | 19.72 |  | 19 | 26.76 |
| McLean University |  | 5 | 7.04 |  | 7 | 9.86 |
| Harvard University |  | 37 | 52.11 |  |  |  |

GM variables were harmonised by ComBat, maintaining biological variations related to diagnosis, sex, and age [[13–15]](https://paperpile.com/c/iqLHPy/OOaGv+zCIJb+R7UWj). The effects of ComBat on our data were evaluated by differences in HCs among imaging sites based on analysis of covariance (Figure S3). The harmonisation process decreased the F-values of most GM variables, largely eliminating significant differences (*p* < 0.05) between scanners. Even after harmonisation, left cuneus and insula thickness still showed a significant difference, but these differences were not significant with Bonferroni correction (*p* < 0.05 / [68 GM regions]).

Multi-site diffusion data were harmonised using two approaches, ComBat and the method utilising rotation invariant spherical harmonics (RISH) features to determine the optimal harmonisation technique [[16, 17]](https://paperpile.com/c/iqLHPy/uya25+FbSG6). For harmonisation with RISH, a minimum of 16 well-matched healthy controls from each imaging site should be recruited, but it is important to note that an insufficient number of subjects or subjects with mismatched ages were recruited from some sites due to the restricted number of subjects (Table S4). For evaluation of the effectiveness of these techniques, the differences between sites for HCs were calculated before and after harmonisation (Figure S4). Although both methods reduced differences in HCs between imaging sites, RISH-based harmonisation exhibited a significant difference in MD with Bonferroni correction (*p* < 0.05 / [44 WM tracts]). As a result, we decided to use the data harmonised by ComBat.

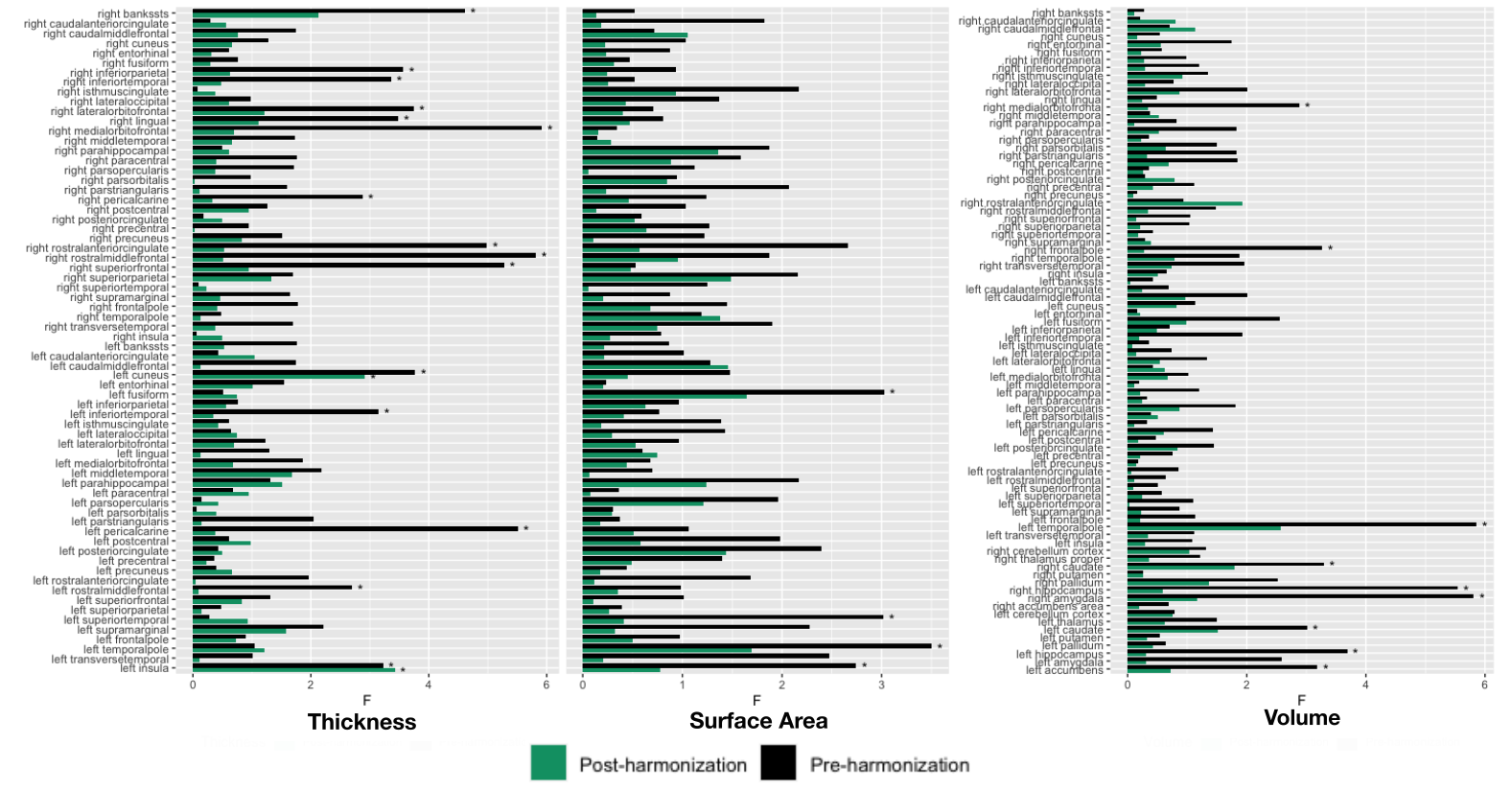

##### Figure S3. Harmonisation on GM variables.

Key: The differences in GM variables among scanners in HCs were assessed using Analysis of Covariance (ANCOVA) with age and sex as covariates. F-values decreased after harmonisation in almost all variables. Asterisks represent significant differences between scanners (*p* < 0.05) largely eliminated by the harmonisation process except for left cuneus thickness and left insula thickness, which was not significant with Bonferroni-correction (*p* < 0.05 / [68 GM regions]).

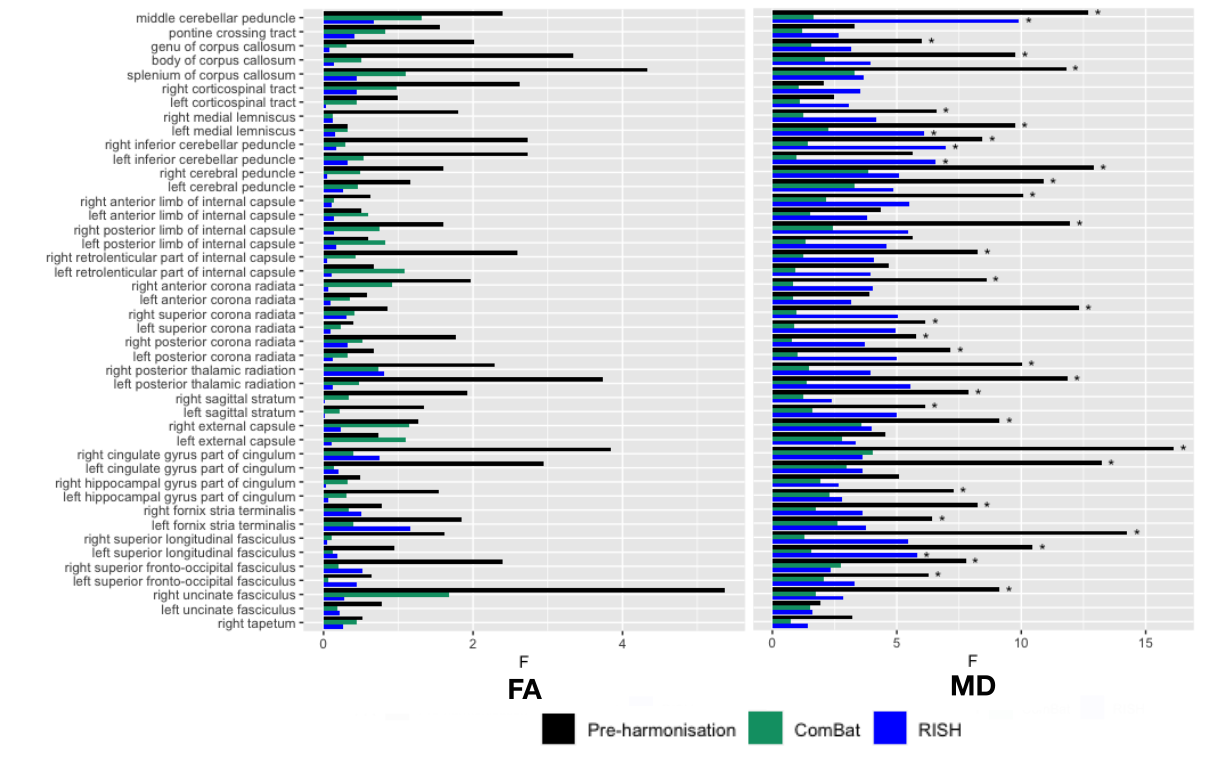

##### Figure S4. Harmonisation on WM variables.

Key: The differences in WM variables between scanners in HCs were assessed using Analysis of Covariance (ANCOVA) with age and sex as covariates. The F-value decreased with both harmonisation techniques in almost all variables. Asterisks represent the significant differences between scanners with Bonferroni-corrected (*p* < 0.05 / [44 WM tracts]). Data harmonised by ComBat did not show significant differences in every variable, while data by RISH showed a significant difference in the MD of some WM tracts.

##### Table S4. Age and sex of controls recruited for harmonisation with RISH features

| Target site name | Age and sex of HCs (Reference site (IU)/**Target site**) |
| --- | --- |
| BWH | 23.82+/-2.34 (SD), 10M, 6F  **24.24+/-1.79 (SD), 10M, 6F** |
| MH | 22.68+/-2.06 (SD), 4M, 1F  **22.55+/-1.98 (SD), 4M, 1F** |
| HU | 22.33+/-2.61 (SD), 9M, 7F  **21.28+/-0.34 (SD), 9M, 7F** |

Key: Comparing the age and sex of HCs selected for harmonisation with rotation invariant spherical harmonics (RISH) features between the reference site (IU) and each target site. IU = Indiana University, BWH = Brigham and Women's Hospital, MH = McLean Hospital, HU = Harvard University.

#### Tractography

Multi-Shell Multi-Tissue Constrained Spherical Deconvolution calculated Fiber Orientation Densities (FOD) to demonstrate diffusion signals' directions based on different tissue types' estimated response functions [[18, 19]](https://paperpile.com/c/iqLHPy/seHVl+JMaQx). Anatomical images were employed to distinguish tissue types to identify the GM-WM boundary [[20]](https://paperpile.com/c/iqLHPy/goMt5). After the co-registration of T1-weighted and diffusion-weighted images, the iFOD2 probabilistic tractography algorithm generated 10 million streamlines per subject, with each voxel on the GM-WM boundary serving as a seed [[21]](https://paperpile.com/c/iqLHPy/LqdpO). The Anatomically Constrained Tractography option was chosen to confine the tractography within WM. An MRtrix command, 'tcksift2', modified the streamlines to match the whole-brain tractography and fibre density at the voxel level, and the total weight of streamlines connecting specific brain regions was calculated based on the parcellation of the DK atlas [[22, 23]](https://paperpile.com/c/iqLHPy/AABrn+luucW).

#### Statistical analyses

*Univariate analysis for demographic, clinical, cognitive, and GM/WM data*

Normality was tested based on the Shapiro-Wilk test, and a test method was selected from the Mann-Whitney U test, chi-square test or ANCOVA based on data type and presence of covariates.

The differences between groups in cognitive test results were evaluated using univariate analysis of covariance (ANCOVA) with sex and age as covariates. Group differences in GM variables were evaluated using ANCOVA with intracranial volume, age, and sex as covariates. Group differences in WM variables were evaluated using ANCOVA with age, sex, and in-scanner absolute and relative head motion as covariates. Bonferroni correction was applied for multiple cognitive, GM, and WM variables comparisons.

#### Multiblock partial least squares correlation analyses (MB-PLS-C)

MB-PLS-C model identifies latent patterns of maximal covariance between disjoint data blocks and between data and covariate blocks. The full details of MB-PLS-C were published [[24]](https://paperpile.com/c/iqLHPy/YHH2b).

WM and GM variables and their covariates were stored in a matrix denoted $X$, $Y$, $C_{X}$, and $C_{Y}$. Each block corresponds to a subject in the row and a variable in the column: $I_{n}\times J_{s}$, $s \in\{X,Y,C_{X},C_{Y}\}$, $n \in\{1,...,N\}$, where $I_{n}$ is the number of subjects in a group, $N$ is the number of groups, and $J_{s}$ is the number of variables of data blocks and covariate blocks. Covariates for the GM variables were age, sex, and intracranial volume, and covariates for the WM variables were age, sex, and in-scanner absolute and relative head motion. A variable of each column was converted into z-scores to enable comparison across variables. The correlation matrices were created between $X$ and $Y$, $X$ and $C_{X}$, $Y$ and $C_{Y}$, and $C_{X}$ and $C_{Y}$, respectively, and combined to create the multiblock cross-product matrix $R$ of size $M\times K$, where $M=N(J_{Y}+J_{C_{X}})$ and $K=J_{X}+J_{C_{Y}}$ (Figure S5). Then, singular value decomposition (SVD) of $R$ was performed,

$R=U\Sigma V^{T},$	(S1)

, where $U$, $\Sigma$, and $V$ are respectively $M\times M$, $M\times K$, and $K\times K$ matrices. $\Sigma$ is a rectangular matrix of $p$ rows and $p$ columns with singular values, $\sigma_{1}>\sigma_{2}>...>\sigma_{p},$on the diagonal ($p =min(M,K)$). Each column of matrix $U$ represents the salience of $Y$ and $C_{X}$ in each LV, and each column of matrix $V$ represents the salience of $X$ and $C_{Y}$ in each LV.

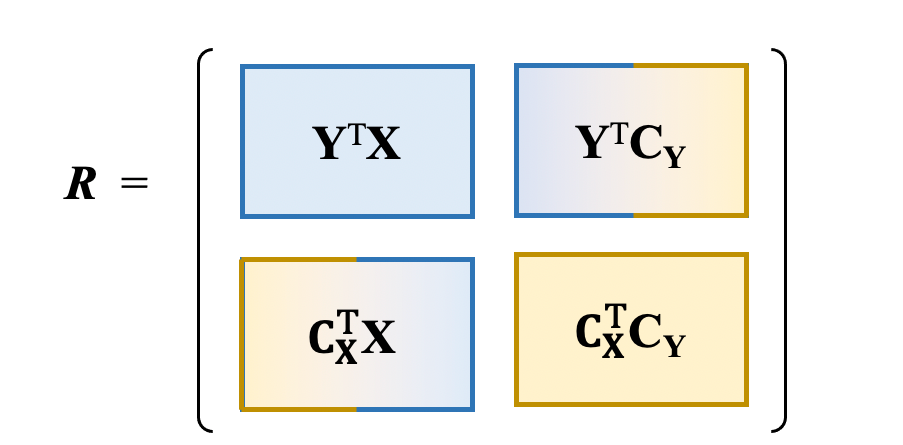

##### Figure S5: The input cross-correlation matrix for MB-PLS-C analysis.

Key: The figure is adapted from Syeda et al. (2022)[[24]](https://paperpile.com/c/iqLHPy/YHH2b).

The latent variable of each LV is computed by multiplying $Y$ and $C_{X}$ by each column of matrix $U$ ($U_{Y}$, $U_{C_{X}}$) and $X$ and $C_{Y}$ by each column of matrix $V$ ($V_{X}$, $V_{C_{Y}}$).

$L_{X}=XV_{X}, L_{Y}=YU_{Y}, L_{C_{X}}=C_{X}U_{C_{X}}, L_{C_{Y}}=C_{Y}V_{C_{Y}},$	(S2)

The definition of LV-specific percent sum-of-squares crossblock covariance is as follows:

$\xi_{i}=\frac{\sigma_{i}^{2}}{\sum_{k=1}^{p} \sigma_{k}^{2}}$,	(S3)

An LV explaining more than 10% of the sum-of-squares crossblock was interpreted as reflecting an underlying characteristic of the data, while an LV explaining less than 10% was regarded as noise.

*Permutation tests*

Permutation tests assessed the statistical significance of the model (omnibus test) and each LV. The order of subjects was randomly synchronously rearranged in $X$, $C_{X}$, and $C_{Y}$, and $R$ blocks were created for singular value decomposition, which was repeated 10 000 times. The significance of the model was assessed by counting the times when the sum of the singular values was equal to or greater than the sum of unpermuted singular values, and the model would be considered significant if the ratio of the times was less than 0.05. Similarly, the significance of LVs was assessed by counting the times when the permuted singular value corresponding to an LV was equal to or greater than the unpermuted singular value, and the LV would be considered significant if the ratio of the times was less than 0.05.

*Bootstrap estimation of confidence intervals*

Bootstrapping was repeated 10 000 times to compute confidence intervals for saliences. $R$ block was created based on resampled data and covariate blocks using random sampling with replacement. Singular value decomposition was performed on $R$ block of each iteration to calculate the saliences in each trial. 95% confidence intervals were calculated from the distribution of saliences.

*Cross-validation*

Monte Carlo cross-validation was used to evaluate the generalisability of the salience pattern. The dataset was randomly partitioned into training and test sets, and the training set was randomly divided into 100 sets of train-validation sets without replacement. Around 80% of the train-validation set (56 subjects from both groups) was configured as the training set. In each train-validation set, $R$ block was created from the training set for singular value decomposition, and patterns of structure and cognition saliences were calculated with permutation test and bootstrapping. The generalisability of the salience patterns was assessed based on the correlation coefficients of the GM-WM latent variables.

*Out-of-sample data*

Out-of-sample data was used to assess the generalisability of the salience pattern. As out-of-sample data, T1- and diffusion-weighted MRI scans of recent-onset psychosis individuals collected in the Melbourne Health and Austin Health were used. The study's data collection was approved by the Melbourne Health and Austin Health ethics committees. The details of acquisition and processing for T1-weighted images are provided in Di Biase et al. (2017)[[25]](https://paperpile.com/c/iqLHPy/Vpe1Q). The diffusion-weighted images were preprocessed with a pipeline that included EPI distortion, eddy current, and motion correction with FSL (v6.0). WM parameters were computed using tract-based spatial statistics analysis (TBSS) for WM tracts of the Johns Hopkins University (JHU) white matter tract atlas [[12]](https://paperpile.com/c/iqLHPy/fjHQC). Subject information is shown in Table S5. The generalisability of the salience patterns was assessed based on the correlation coefficients of the GM-WM latent variables calculated by multiplying the values from the out-of-sample and the salience calculated from the original datasets.

##### Table S5. Subject information of out-of-sample

| Variables |  | Healthy Controls  (*n* = 25) | |  | Recent-Onset Psychosis Individuals  (*n* = 26) | |
| --- | --- | --- | --- | --- | --- | --- |
|  |  | **n** | **%** |  | **n** | **%** |
| Sex, male |  | 16 | 64.00 |  | 19 | 73.08 |
|  |  | **Mean** | **SD** |  | **Mean** | **SD** |
| Age |  | 21.92 | 1.87 |  | 21.09 | 1.93 |
| Duration of illness |  |  |  |  | 1.82 | 0.83 |
| Age of symptom onset |  |  |  |  | 19.27 | 2.14 |
| Symptoms |  |  |  |  |  |  |
| General pathology (BPRS total) |  |  |  |  | 38.92 | 9.29 |
| Positive symptoms (BPRS subscore) |  |  |  |  | 12.28 | 4.36 |
| Negative symptoms (SANS) |  |  |  |  | 17.80 | 10.08 |
| General functioning (SOFAS) |  |  |  |  | 55.27 | 8.98 |

#### Normalised between-group structural salience difference (NSSD)

To evaluate group differences in GM patterns between HCs and ROP individuals, the normalised between-group structural salience difference (NSSD) for the $i$-th LV and $j$-th GM variable is calculated using the following equation.

$$\kappa_{ij} =\frac{\left| u_{ij,HC} \right|-\left| u_{ij,ROP} \right|}{s_{pool}}$$

$u_{ij,HC}$ and $u_{ij,ROP}$ represent the salience for the $i$-th LV and the $j$-th GM variable of each group. $s_{pool}$ is the expected variation which is calculated by pooling the confidence intervals of two groups using the following equation.

$$s_{pool}=\sqrt{\frac{\left( n_{HC}-1 \right)s_{ij,HC}^{2}+\left( n_{ROP}-1 \right)s_{ij,ROP}^{2}}{n_{HC}+n_{ROP}-2}}$$

$n_{HC}$ and $n_{ROP}$ are the number of subjects in each group, and $s_{ij,HC}$ and $s_{ij,ROP}$ are the sum of the lower and upper confidence intervals of each group.

A positive NSSD value indicates the stronger contribution of the $j$-th HC salience to the $i$-th GM LV than the one of the $j$-th ROP salience, while a negative NSSD value means the stronger contribution of the $j$-th ROP salience than the one of the $j$-th HC salience.

#### Sample size and salience convergence

For examination of whether the values of saliences converged with increasing sample size, the salience values were calculated when subjects were randomly selected with a different sample size. The absolute difference in each salience between N+1 and N samples was calculated, and the mean of the differences was graphed to see their trajectories with increasing sample size. If the mean has converged enough as the sample size increases, the salience could be evaluated as stable with a sufficient sample size.

#### Comparison between latent cortical patterns and allometric scaling maps

We compared the cortical salience patterns of two groups with the allometric scaling maps using the NIH data[[26]](https://paperpile.com/c/iqLHPy/ENM7I). All the cortical volume maps were transformed to the *'fsaverage'* coordinate system at a density of 41K vertices per hemisphere using the Neuromaps toolbox in Python. The allometric scaling maps were parcellated based on the Desikan-Killiany atlas using MATLAB, and all parcellated maps were visually inspected for accuracy. As the Desikan-Killiany atlas in the Neuromaps toolbox was not available, pearson correlation coefficients between the salience patterns of the HC group and the ROP group and the allometric scaling patterns were calculated using vertex data in MATLAB (*'corr'* function).

### RESULTS

#### Case-control differences in GM and WM

Group differences in GM variables controlling for age, sex and total intracranial volume is shown in Figure S6. Group differences in WM variables controlling for age, sex, and in-scanner absolute and relative head motion are shown in Figure S7.

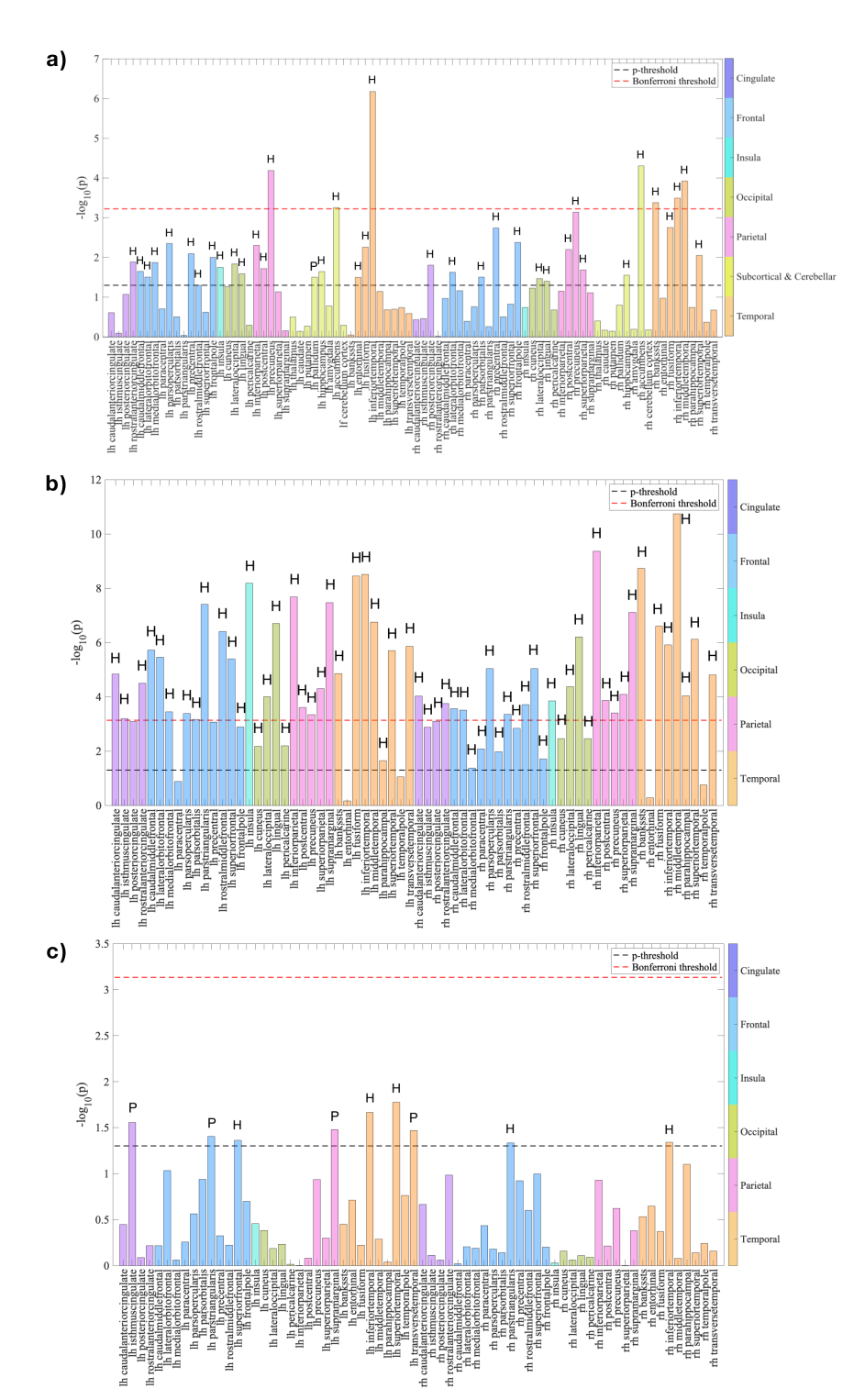

##### Figure S6. Group differences in GM variables between recent onset psychosis and healthy controls.

Key: a) GM volume, b) GM thickness, c) GM surface area. The significance of log-transformed *p*-values was assessed by -log_10_ (0.05) (black line), and the Bonferroni corrected threshold, -log_10_ (0.05 / [84 GM regions]) in a) and -log_10_ (0.05 / [68 GM regions]) in b) and c) (red line). The letters “H” or “P” at the end of the bar graph, respectively, exhibit healthy controls (“H”) and ROP individuals (“P”) with a higher value. Following regions represented statistically significant results with the Bonferroni-corrected p-value threshold: a) the bilateral nuclei accumbens, inferior temporal gyrus, left postcentral gyrus, right banks of the superior temporal sulcus, middle temporal gyrus, b) the bilateral caudal anterior cingulate gyrus, rostral anterior cingulate gyrus, caudal middle frontal gyrus, lateral orbitofrontal gyrus, pars opercularis, pars triangularis, rostral middle frontal gyrus, superior frontal gyrus, insula, lateral occipital gyrus, lingual, inferior parietal gyrus, postcentral gyrus, precuneus, superior parietal gyrus, supramarginal gyrus, banks of the superior temporal sulcus, fusiform, inferior temporal gyrus, middle temporal gyrus, superior temporal gyrus, transverse temporal gyrus, left isthmus cingulate gyrus, medial orbitofrontal gyrus, pars orbitalis, right parahippocampal gyrus, c) the bilateral pars triangularis, inferior temporal gyrus, left isthmus cingulate gyrus, superior frontal gyrus, supramarginal gyrus, superior temporal gyrus, transverse temporal gyrus.

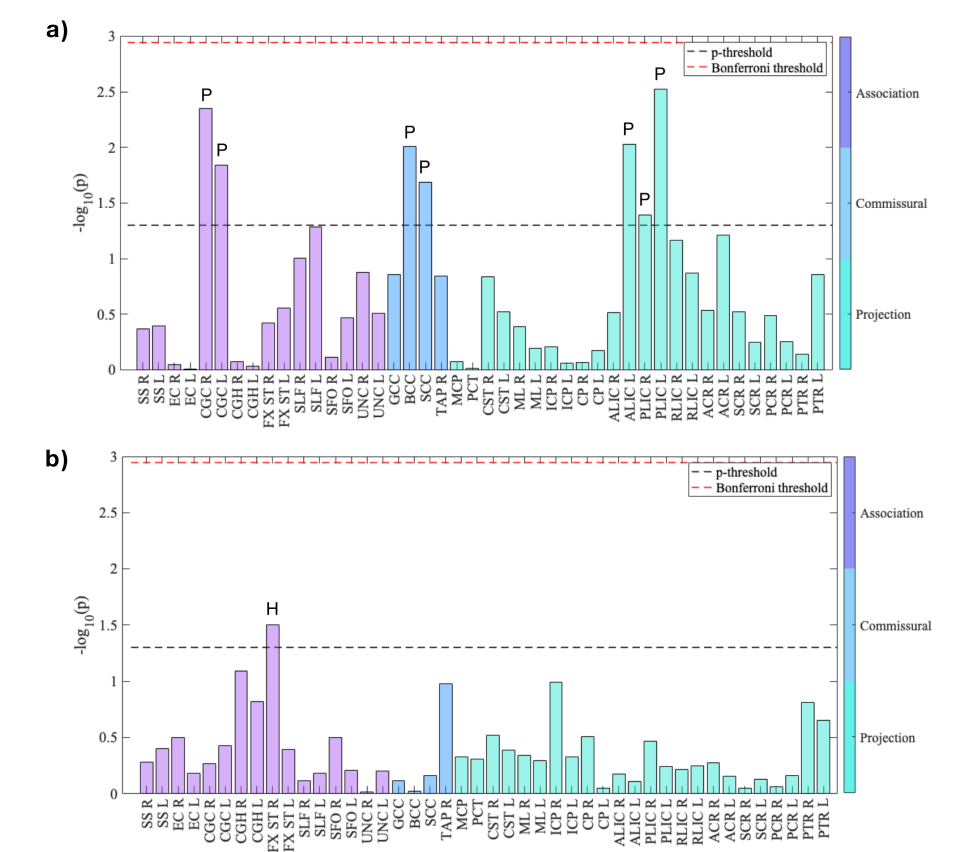

##### Figure S7. Group differences in WM variables between recent onset psychosis and healthy controls.

Key: The significance of log-transformed *p*-values is assessed by -log_10_ (0.05) (black line), and the Bonferroni corrected threshold, -log_10_ (0.05 / [44 WM tracts]) (red line). a) FA, b) MD. The letters "H" or "P" at the end of the bar graph, respectively, denote healthy controls ("H") and ROP individuals ("P") with higher values. SS: Sagittal stratum, EC: External capsule, CGC: Cingulum (cingulate gyrus), CGH: Cingulum (hippocampus), FX/ST: Fornix/Stria terminalis, SLF: Superior longitudinal fasciculus, SFO: Superior fronto-occipital fasciculus, UNC: Uncinate fasciculus, GCC: Genu of corpus callosum, BCC: Body of corpus callosum, SCC: Splenium of corpus callosum, TAP: Tapatum, MCP: Middle cerebellar peduncle, PCT: Pontine crossing tract, CST: Corticospinal tract, ML: Medial lemniscus, ICP: Inferior cerebellar peduncle, CP: Cerebral peduncle, ALIC: Anterior limb of the internal capsule, PLIC: Posterior limb of the internal capsule, RLIC: Retrolenticular part of the internal capsule, ACR: Anterior corona radiata, SCR: Superior corona radiata, PCR: Posterior corona radiata, PTR: Posterior thalamic radiation

##### Table S6. Group differences in cognitive variables.

| Variables |  | Healthy Controls  (*n* = 71) | |  | ROP individuals  (*n* = 71) | |  | Test statistic  Effect size |
| --- | --- | --- | --- | --- | --- | --- | --- | --- |
|  |  | **Mean** | **SD** |  | **Mean** | **SD** |  |  |
| Working memory |  | 19.6 | 2.7 |  | 16.1 | 3.8 |  | †F = 38.89*, Cohen *d* = 1.062 |
| Episodic memory |  | 0.24 | 0.86 |  | -0.85 | 0.90 |  | †F = 53.93*, Cohen *d* = 1.238 |
| Processing speed |  | 53.1 | 6.6 |  | 45.0 | 8.2 |  | †F = 44.15*, Cohen *d* = 1.088 |
| Word reading skill |  | 6.90 | 2.21 |  | 4.27 | 3.31 |  | †F = 30.04*, Cohen *d* = 0.935 |

Key: * *p*<0.001

† the *F*-statistic report the group effect in analysis of covariance with age and sex as a covariance

#### PLS between GM thickness and WM FA

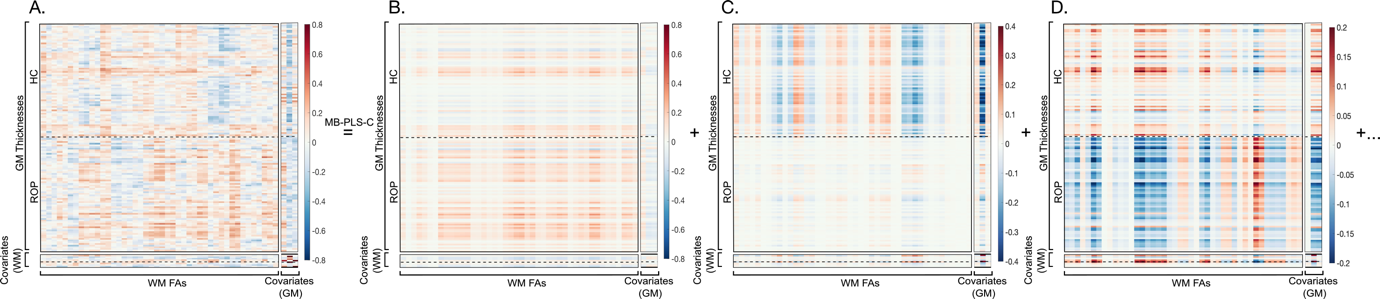

##### Figure S8. Decomposition of the input multi-block cross-correlation matrix by MB-PLS-C between GM thickness and WM FA.

Key: A: input correlation block, B: LV1, C: LV2, D: LV3. Covariates (WM) represent age, in-scanner absolute and relative head motion, and sex by group from the top. Covariates (GM) represent intracranial volume, age, and sex from the left.

##### Table S7. MB-PLS-C latent variables contributing to the relationship between GM thickness and WM FA.

| Latent component | Contribution to 𝜎_𝑖_* (%) | | | | Sum-of-squares covariance, 𝜉𝑖* (%) | *p*-value |
| --- | --- | --- | --- | --- | --- | --- |
|  | block 1 (*𝐿_FA_^𝑇^𝐿_TH_*) | block 2 (*𝐿_FA_^𝑇^𝐿_CFA_*) | block 3 (*𝐿_CTH_^𝑇^𝐿_TH_*) | block 4 (*𝐿_CTH_^𝑇^𝐿_CFA_*) |  |  |
| LV1 | 90.63 | 5.78 | 3.44 | 0.14 | 34.30 | 0.33 |
| LV2 | 66.36 | 1.28 | 28.24 | 4.12 | 16.92 | 0.040 |
| LV3 | 84.22 | 3.86 | 9.41 | 2.50 | 12.38 | 0.003 |

Key: The values represent the block-wise contributions of latent variables to the singular values of the significant latent components derived from the MB-PLS-C between GM thickness and WM FA

^*^𝜎_𝑖_ and 𝜉_𝑖_ are the singular values and percent sum-of-squares crossblock covariances corresponding to the *i*th significant LV, 𝑖∈{1,2,3}.

*L_FA_*=latent variables of FA; *L_TH_*=latent variables of thickness; *L_CFA_*=latent variables of covariates of FA; *L_CTH_*=latent variables of covariates of thickness

#### PLS between GM surface area and WM FA

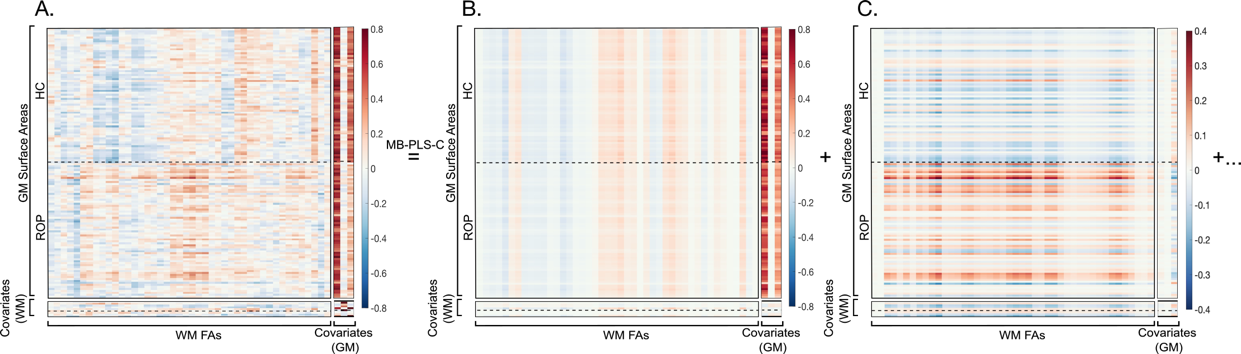

##### Figure S9. Decomposition of the input multi-block cross-correlation by MB-PLS-C between GM surface area and WM FA.

Key: A: input correlation block, B: LV1, C: LV2. Covariates (WM) represent age, in-scanner absolute and relative head motion, and sex by group from the top. Covariates (GM) represent intracranial volume, age, and sex from the left.

##### Table S8. MB-PLS-C latent variables contributing to the relationship between GM surface area and WM FA.

| Latent component | Contribution to 𝜎_𝑖_* (%) | | | | Sum-of-squares covariance, 𝜉𝑖* (%) | *p*-value |
| --- | --- | --- | --- | --- | --- | --- |
|  | block 1 (𝐿*_FA_*^𝑇^𝐿*_SA_*) | block 2 (*𝐿_FA_^𝑇^𝐿_CFA_*) | block 3 (*𝐿_CSA_^𝑇^𝐿_SA_*) | block 4 (*𝐿_CSA_^𝑇^𝐿_CFA_*) |  |  |
| LV1 | 29.67 | 0.61 | 67.86 | 1.86 | 53.21 | 0.013 |
| LV2 | 91.13 | 6.81 | 1.86 | 0.20 | 18.97 | 0.0003 |

Key: The values represent the block-wise contributions of latent variables to the singular values of the significant latent components derived from the MB-PLS-C between GM surface area and WM FA. ^*^𝜎_𝑖_ and 𝜉_𝑖_ are the singular values and percent sum-of-squares crossblock covariances corresponding to the *i*th significant LV, 𝑖∈{1,2}. *L_FA_*=latent variables of FA; *L_SA_*=latent variables of surface area; *L_CFA_*=latent variables of covariates of FA; *L_CSA_*=latent variables of covariates of surface area

#### PLS between GM volume and WM FA

The MB-PLS-C analyses between GM volume and WM FA were significant (omnibus test *p* < 0.0005). The input multi-block cross-correlation matrix was decomposed into multi-block cross-correlation matrices in each latent dimension by MB-PLS-C (Figure S10). LV1 and LV2 were significant and explained 70.76% of sum-of-squares covariance (Table S9).

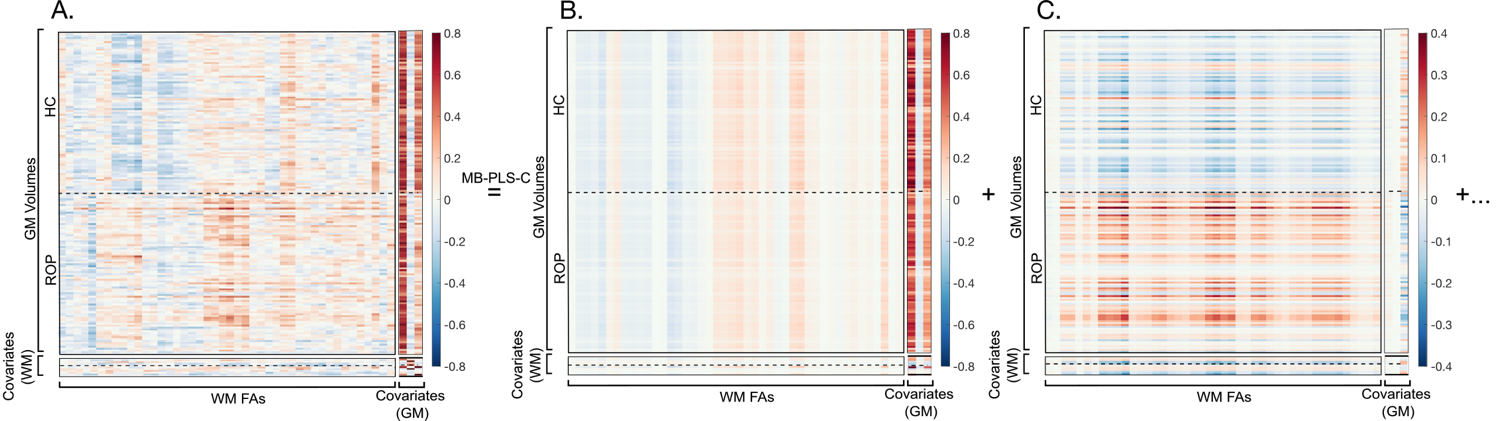

##### Figure S10. Decomposition of the input multi-block cross-correlation by MB-PLS-C between GM volume and WM FA.

Key: A: input correlation block, B: LV1, C: LV2). Covariates (WM) represent age, in-scanner absolute and relative head motion, and sex by group from the top. Covariates (GM) represent intracranial volume, age, and sex from the left.

##### Table S9. MB-PLS-C latent variables contributing to the relationship between GM volume and WM FA.

| Latent component | Contribution to 𝜎_𝑖_* (%) | | | | Sum-of-squares covariance, 𝜉𝑖* (%) | *p*-value |
| --- | --- | --- | --- | --- | --- | --- |
|  | block 1 (𝐿*_FA_*^𝑇^𝐿*_VOL_*) | block 2 (*𝐿_FA_^𝑇^𝐿_CFA_*) | block 3 (*𝐿_CVOL_^𝑇^𝐿_VOL_*) | block 4 (*𝐿_CVOL_^𝑇^𝐿_CFA_*) |  |  |
| LV1 | 30.49 | 0.51 | 67.34 | 1.66 | 50.84 | 0.016 |
| LV2 | 91.47 | 5.27 | 3.01 | 0.24 | 19.91 | 0.0001 |

Key: ^*^𝜎_𝑖_ and 𝜉_𝑖_ are the singular value and percent sum-of-squares crossblock covariance corresponding to the *i*th significant LV, 𝑖∈{1,2}. *L_FA_*=latent variables of FA; *L_VOL_*=latent variables of volume; *L_CFA_*=latent variables of covariates of FA; *L_CVOL_*=latent variables of covariates of volume

*First latent variable*

LV1 explained 50.85% of sum-of-squares crossblock covariance (*p* < 0.0005), and both groups showed similar correlations. In the covariate blocks, total intracranial volume and male sex were positively correlated with GM volume.

All significant saliences of GM volume in LV1 were positively weighted and exhibited a shared GM pattern between the groups, with the HC group demonstrating a stronger mapping to LV1 than the ROP group (Figure S11A). The ROP group presented the largest saliences in temporal and subcortical regions: the left thalamus and pallidum and right thalamus and middle and inferior temporal gyri. The HC group demonstrated the largest saliences in temporal and subcortical regions and insula: the left insula and hippocampus and right middle, superior, and inferior temporal gyri. The largest positive saliences of WM FA were observed in the right uncinate fasciculus, left external capsule, and left superior corona radiata, whereas the largest negative saliences were in the bilateral anterior limb of the internal capsule and splenium of corpus callosum (Figure S11B).

In the LV-specific NSSD metric explaining group differences in saliences of GM regions, the HC group displayed the most robust NSSD metric in the bilateral hippocampal gyrus, left entorhinal cortex, and right superior temporal gyrus, pars orbitalis, and pars triangularis. The ROP group exhibited the most robust NSSD metric in the bilateral lingual gyrus, left cuneus, pericalcarine cortex, and nucleus accumbens, and right superior parietal gyrus (Figure S11C). In the training sample (n=142), the control group displayed a slightly greater positive correlation between latent variables of GM volume and WM FA (HC: *r* = 0.50, *p* < 0.001, ROP: *r* = 0.43, *p* < 0.001). Out-of-sample data (n=51) demonstrated a high correlation in ROP individuals, but a significant correlation was not observed in the cross-validation data (n=112 in training set, n=30 in test set) (Figure S11D). After 50 samples for each group, the GM and WM saliences showed convergence when the sample size reached 49 per group (Figure S11E).

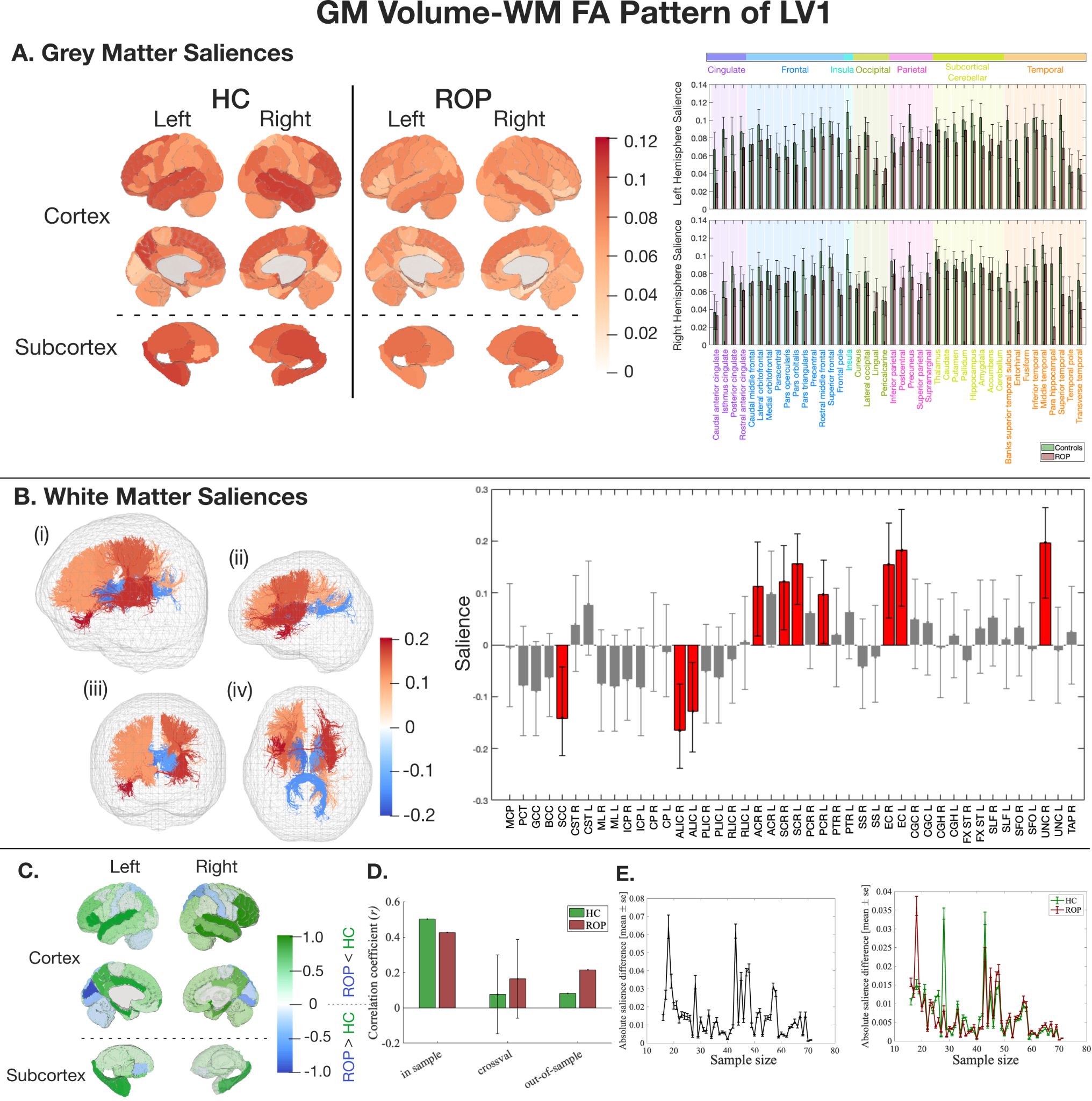

##### Figure S11. The latent pattern of LV1 between GM volume and WM FA derived from MB-PLS-C.

Key: A) GM salience by group in HC and ROP individuals. GM saliences of HC (green) and ROP (red) with 95% confidence interval (black lines) and color-coded lobe information. All the reliable saliences were positive. B) WM saliences shared between groups. WM tracts represented in tractography are color-coded based on the salience intensity ((i) overall view (ii) from the left (iii) from the front (iv) from the bottom). The bar plot shows the salience of each WM tract with 95% confidence intervals (black lines). C) Normalised regional salience-differences. A positive score (green) shows a salience is larger in HC, and a negative score (blue) shows a salience is larger in ROP. D) A correlation coefficient between LV1 of GM and WM in the training and the test samples, and a correlation coefficient and its standard deviation from Monte Carlo cross-validation. While the cross-validation data did not show a significant correlation, out-of-sample data showed generalisability in the ROP group*.* E) The impact of sample size on salience intensity. Figures on the left and right depict differences in WM saliences and GM saliences, respectively. The GM-WM pattern converged with 50 samples from each group.

ACR=anterior corona radiata; ALIC=anterior limb of the internal capsule; BCC=body of corpus callosum; CGC=cingulum (cingulate gyrus); CGH=cingulum (hippocampal portion); CP=cerebral peduncle; CR=corona radiata; CST=corticospinal tract; EC=external capsule; FX ST=fornix (cres) / stria terminalis; GCC=genu of corpus callosum; ICP=interior cerebellar peduncle; ML=medial lemniscus; MCP=middle cerebellar peduncle; PCT=pontine crossing tract; PCR=posterior corona radiata; PLIC=posterior limb of the internal capsule; PTR=posterior thalamic radiation; RLIC=retrolenticular part of the internal capsule; SCC=splenium of corpus callosum; SCR=superior corona radiata; SFO=superior fronto-occipital fasciculus; SLF=superior longitudinal fasciculus; SS=sagittal stratum; TAP=tapetum; UNC=uncinate fasciculus.

*Second latent variable*

LV2 explained 19.91% of sum-of-squares crossblock covariance (*p* = 0.0001), and the HC group displayed negative correlations, whereas the ROP demonstrated positive correlations.

Significant saliences of GM volume demonstrated differential GM patterns between the ROP and HC groups (Figure S12A). The HC group demonstrated the largest saliences in frontal and subcortical regions: the left nucleus accumbens, thalamus, and frontal pole and right pars opercularis and caudal middle frontal gyrus. The ROP group presented the largest saliences in frontal and parietal regions: the bilateral precentral gyrus, left paracentral and caudal middle frontal gyri, and right post central gyrus. For WM FA, the largest negative saliences were present in the bilateral inferior cerebellar peduncle, superior corona radiata, left posterior corona radiata, and right medial lemniscus (Figure S12B).

The HC group exhibited the most robust NSSD metric in the left posterior cingulate cortex and right pars opecularis, putamen, and ligual gyrus. The ROP group displayed the most robust NSSD metric in the bilateral superior parietal gyrus, left precentral gyrus and cerebellar cortex, and right superior frontal and caudal middle frontal gyri (Figure S12C). In the training sample (n’s as above), the ROP group displayed a greater positive correlation between latent variables of GM GM volume and WM FA (HC: *r* = 0.25, *p* = 0.038, ROP: *r* = 0.38, *p* = 0.0011). A similar correlation was observed in out-of-sample data (n’s as above) in healthy controls, while a significant correlation was not observed in the cross-validation data (n’s as above) (Figure S12D). After 50 samples per group, the GM and WM saliences showed convergence when the sample size reached 49 per group (Figure S12E).

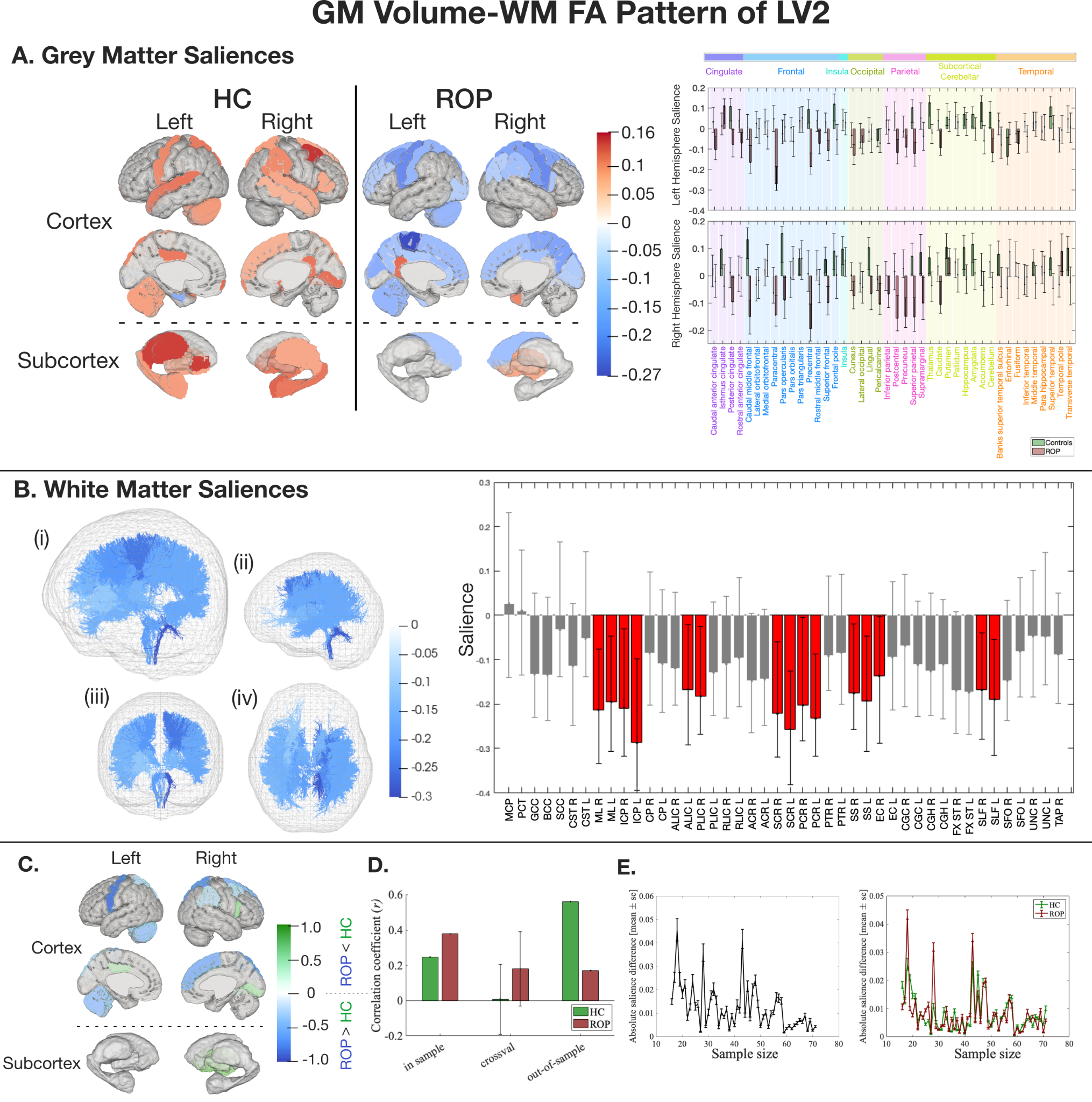

##### Figure S12. The latent pattern of LV2 between GM volume and WM FA derived from MB-PLS-C.

Key: A) GM salience by group in HC and ROP individuals. GM saliences of HC (green) and ROP (red) with 95% confidence interval (black lines) and color-coded lobe information. The most of the reliable saliences in HC were positive, while those in ROP were negative. B) WM saliences shared between groups. WM tracts represented in tractography are colour-coded based on the salience intensity ((i) overall view (ii) from the left (iii) from the front (iv) from the bottom). The bar plot shows the salience of each WM tract with 95% confidence intervals (black lines). C) Normalised regional salience-differences. A positive score (green) shows a salience is larger in HC, and a negative score (blue) shows a salience is larger in ROP. D) A correlation coefficient between LV2 of GM and WM in the training and the test samples, and a correlation coefficient and its standard deviation from Monte Carlo cross-validation. A significant correlation was not seen in the cross-validation data, although generalisability was seen in out-of-sample data in healthy controls. E) The impact of sample size on salience intensity. Figures on the left and right depict differences in WM saliences and GM saliences, respectively. After 50 samples for each group, the GM and WM saliences illustrated a tendency to converge.

ACR=anterior corona radiata; ALIC=anterior limb of the internal capsule; BCC=body of corpus callosum; CGC=cingulum (cingulate gyrus); CGH=cingulum (hippocampal portion); CP=cerebral peduncle; CR=corona radiata; CST=corticospinal tract; EC=external capsule; FX ST=fornix (cres) / stria terminalis; GCC=genu of corpus callosum; ICP=interior cerebellar peduncle; ML=medial lemniscus; MCP=middle cerebellar peduncle; PCT=pontine crossing tract; PCR=posterior corona radiata; PLIC=posterior limb of the internal capsule; PTR=posterior thalamic radiation; RLIC=retrolenticular part of the internal capsule; SCC=splenium of corpus callosum; SCR=superior corona radiata; SFO=superior fronto-occipital fasciculus; SLF=superior longitudinal fasciculus; SS=sagittal stratum; TAP=tapetum; UNC=uncinate fasciculus.

#### PLS between GM volume and WM MD

The MB-PLS-C analyses between GM volume and WM MD were significant (omnibus test *p* < 0.0005). The input multi-block cross-correlation matrix was decomposed into multi-block cross-correlation matrices in each latent dimension by MB-PLS-C (Figure S13). LV1 and LV2 were significant and explained 85.93% of sum-of-squares covariance (Table S10).

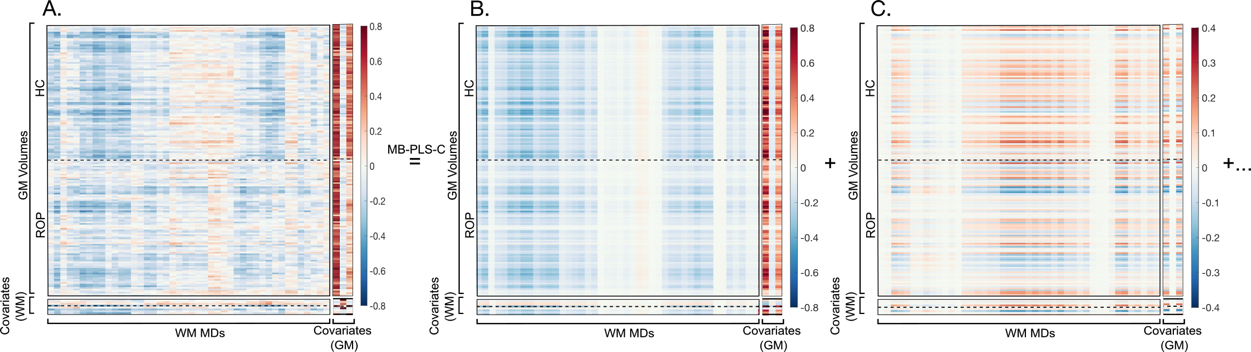

##### Figure S13. Decomposition of the input multi-block cross-correlation matrix by MB-PLS-C between GM volume and WM MD.

Key: A: input correlation block, B: LV1, C: LV2. Covariates (WM) represent age, in-scanner absolute and relative head motion, and sex by group from the top. Covariates (GM) represent intracranial volume, age, and sex from the left.

##### Table S10. MB-PLS-C latent variables contributing to the relationship between GM volume and WM MD.

| Latent component | Contribution to 𝜎_𝑖_* (%) | | | | Sum-of-squares covariance, 𝜉𝑖* (%) | *p*-value |
| --- | --- | --- | --- | --- | --- | --- |
|  | block 1 (𝐿*_MD_*^𝑇^𝐿*_VOL_*) | block 2 (*𝐿_MD_^𝑇^𝐿_CMD_*) | block 3 (*𝐿_CVOL_^𝑇^𝐿_VOL_*) | block 4 (*𝐿_CVOL_^𝑇^𝐿_CMD_*) |  |  |
| LV1 | 66.42 | 3.47 | 29.17 | 0.94 | 74.83 | 0.0003 |
| LV2 | 81.79 | 6.34 | 11.66 | 0.21 | 11.10 | 0.026 |

Key: ^*^𝜎_𝑖_ and 𝜉_𝑖_ are the singular value and percent sum-of-squares crossblock covariance corresponding to the *i*th significant LV, 𝑖∈{1,2}.*L_MD_*=latent variables of MD; *L_VOL_*=latent variables of volume; *L_CMD_*=latent variables of covariates of MD; *L_CVOL_*=latent variables of covariates of volume

*First latent variable*

LV1 explained 74.83% of sum-of-squares crossblock covariance (*p* = 0.0003), and both groups demonstrated similar correlations. In the covariate blocks, total intracranial volume and male sex were positively correlated with GM volume.

All significant saliences of GM volume in LV1 were negatively weighted and exhibited a shared GM pattern between groups (Figure S14A). The ROP group exhibited the largest saliences in parietal, temporal, and subcortical regions: the bilateral pallidum, left nucleus accumbens and inferior parietal gyrus, and right thalamus and inferior temporal gyrus. The HC group showed the largest saliences in cingulate and frontal regions: the bilateral lateral orbitofrontal gyrus, left superior frontal and rostral anterior cingulate gyri, and right rostral middle frontal gyrus and pars triangularis. The largest positive saliences of WM MD were observed in the pontine crossing tract, bilateral medial lemniscus and cerebral peduncle, and right corticospinal tract (Figure S14B).

The HC group showed the most robust NSSD metric in the bilateral lateral orbitofrontal gyrus, left superior frontal and medial orbitofrontal gyri, and right parahippocampal gyrus and pars triangularis. The ROP group exhibited the most robust NSSD metric in the left pericalcarine, cuneus, pallidum, and nucleus accumbens and right lingual gyrus and putamen (Figure S14C). In the training sample (n’s as above), the control group displayed a slightly greater positive correlation between latent variables (HC: *r* = 0.39, *p* < 0.001, ROP: *r* = 0.33, *p* = 0.005) and both groups demonstrated a significant correlation in the cross-validation data (n’s as above), whereas out-of-sample data (n’s as above) didn’t demonstrate a similar pattern (Figure S14D). GM and WM saliences exhibited convergence when the sample size reached 42 per group (Figure S14E).

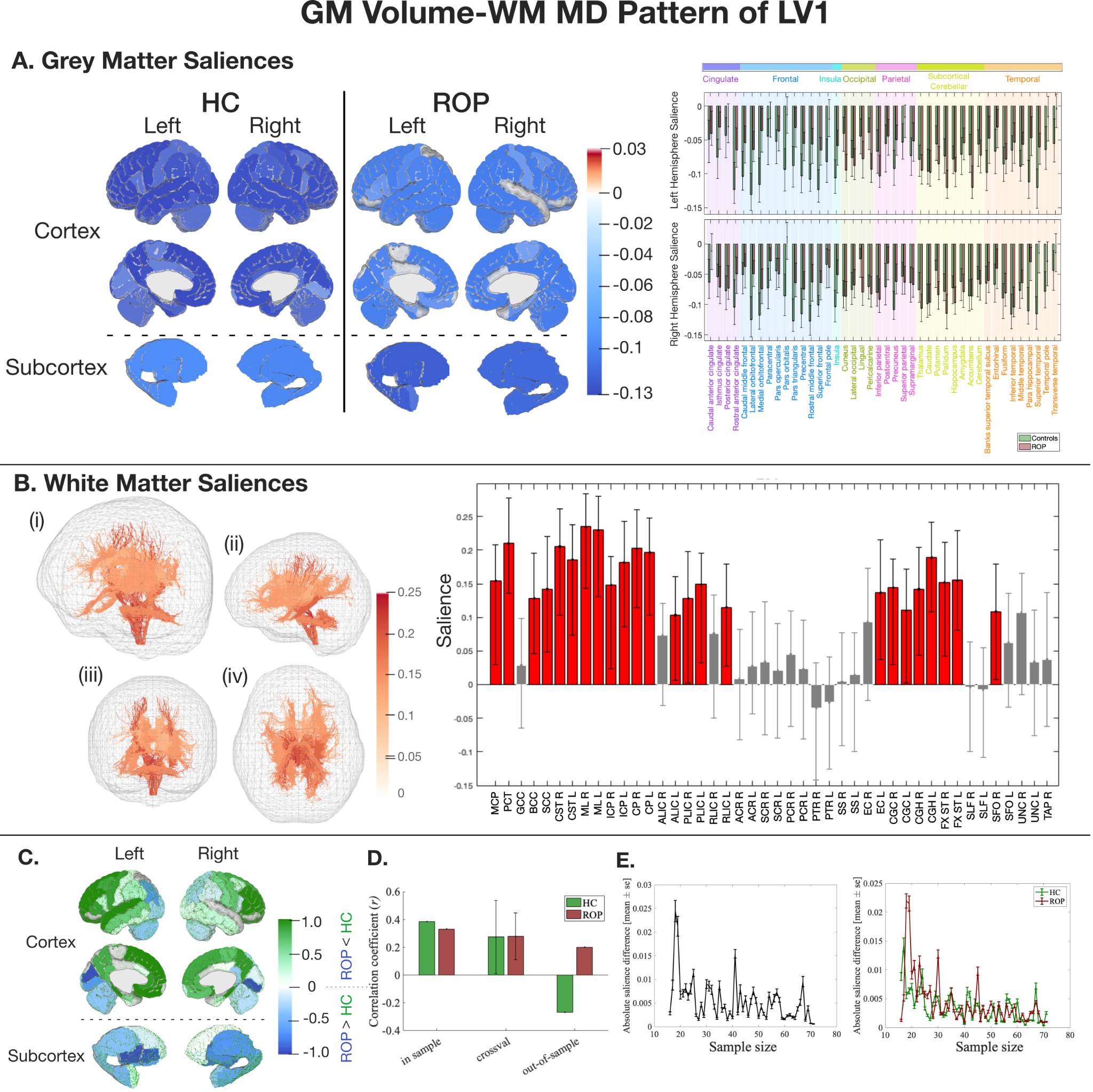

##### Figure S14. The latent pattern of LV1 between GM volume and WM MD derived from MB-PLS-C.

Key: A) GM salience by group in HC and ROP individuals. GM saliences of HC (green) and ROP (red) with 95% confidence interval (black lines) and color-coded lobe information. All the reliable saliences were negative. B) WM saliences shared between groups. WM tracts represented in tractography are colour-coded based on the salience intensity ((i) overall view (ii) from the left (iii) from the front (iv) from the bottom). The bar plot shows the salience of each WM tract with 95% confidence intervals (black lines). C) Normalised regional salience-differences. A positive score (green) shows a salience is larger in HC, and a negative score (blue) shows a salience is larger in ROP. D) A correlation coefficient between LV1 of GM and WM in the training and the test samples, and a correlation coefficient and its standard deviation from Monte Carlo cross-validation. In the cross-validation data, both groups showed significant correlations, but out-of-sample data didn't show a similar pattern. E) The impact of sample size on salience intensity. Figures on the left and right depict differences in WM saliences and GM saliences, respectively. The GM-WM pattern converged with 42 samples from each group.

ACR=anterior corona radiata; ALIC=anterior limb of the internal capsule; BCC=body of corpus callosum; CGC=cingulum (cingulate gyrus); CGH=cingulum (hippocampal portion); CP=cerebral peduncle; CR=corona radiata; CST=corticospinal tract; EC=external capsule; FX ST=fornix (cres) / stria terminalis; GCC=genu of corpus callosum; ICP=interior cerebellar peduncle; ML=medial lemniscus; MCP=middle cerebellar peduncle; PCT=pontine crossing tract; PCR=posterior corona radiata; PLIC=posterior limb of the internal capsule; PTR=posterior thalamic radiation; RLIC=retrolenticular part of the internal capsule; SCC=splenium of corpus callosum; SCR=superior corona radiata; SFO=superior fronto-occipital fasciculus; SLF=superior longitudinal fasciculus; SS=sagittal stratum; TAP=tapetum; UNC=uncinate fasciculus.

*Second latent variable*

LV2 explained 11.10% of sum-of-squares crossblock covariance (*p* = 0.026), and no clear group-specific patterns were observed in correlations of the data block and covariate blocks.

In the HC group, the majority of the significant saliences of GM volume displayed positive correlations, while in the ROP group, approximately half of the significant saliences were positive, and the other half were negative (Figure S15A). The HC group had the largest saliences in cingulate, frontal, occipital, subcortical regions and cerebellum: left posterior cingulate gyrus and cerebellum and right amygdala, hippocampus, lingual gyrus, and pars opercularis. The ROP group had the positively largest saliences in cingulate, frontal, and temporal regions: the left isthmus cingulate gyrus and pars triangularis and right pars orbitalis and temporal pole. In contrast, it had the negatively largest saliences in frontal and occipital regions: the bilateral precentral gyrus and left cuneus and pericalcarine. For WM MD, the largest negative saliences were observed in the bilateral superior longitudinal fasciculus and superior corona radiata, left anterior corona radiata, and right posterior corona radiata (Figure S15B).

The HC group exhibited the most robust NSSD metric in the left amygdala and right lingual, lateral orbitofrontal gyri, and caudal middle frontal gyri. The ROP group displayed the most robust NSSD metric in the left cuneus, middle temporal, inferior temporal, and caudal anterior cingulate gyri and right precentral gyrus (Figure S15C). In the training sample (n’s as above), the ROP group only showed a significant positive correlation between GM volume and WM MD (HC: *r* = 0.23, *p* = 0.054, ROP: *r* = 0.61, *p* < 0.001). The cross-validation data (n’s as above) did not show a significant correlation, but out-of-sample data (n’s as above) from healthy controls demonstrated generalisability (Figure S15D). GM and WM saliences demonstrated convergence when the sample size reached 42 per group (Figure S15E).

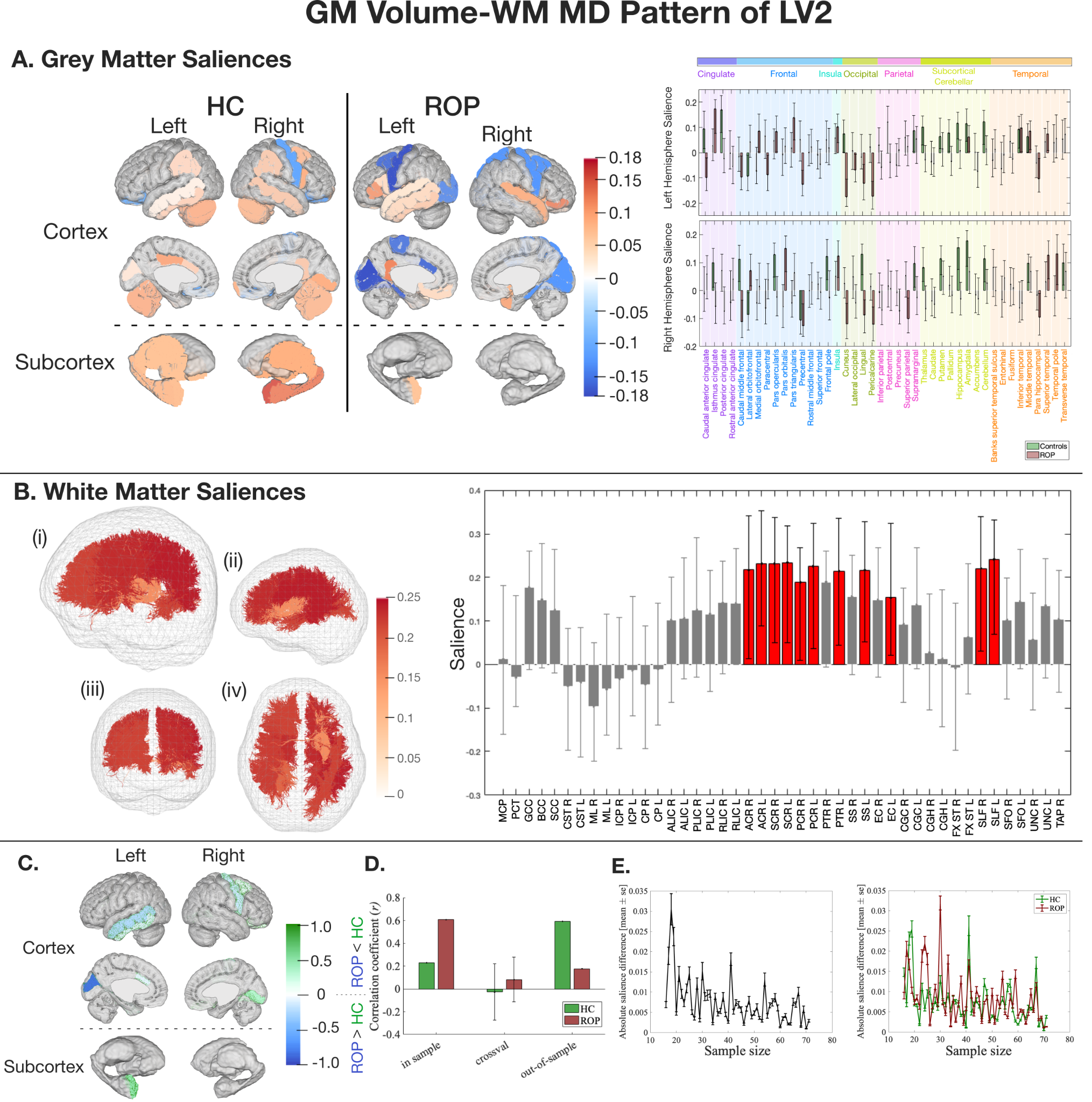

##### Figure S15. The latent pattern of LV2 between GM volume and WM MD derived from MB-PLS-C.

Key: A) GM salience by group in HC and ROP individuals. GM saliences of HC (green) and ROP (red) with 95% confidence interval (black lines) and color-coded lobe information. B) WM saliences shared between groups. WM tracts represented in tractography are colour-coded based on the salience intensity ((i) overall view (ii) from the left (iii) from the front (iv) from the bottom). The bar plot shows the salience of each WM tract with 95% confidence intervals (black lines). C) Normalised regional salience-differences. A positive score (green) shows a salience is larger in HC, and a negative score (blue) shows a salience is larger in ROP. D) A correlation coefficient between LV2 of GM and WM in the training and the test samples, and a correlation coefficient and its standard deviation from Monte Carlo cross-validation*.* Healthy controls' out-of-sample data displayed generalisability, but the cross-validation data did not demonstrate a statistically significant correlation. E) The impact of sample size on salience intensity. Figures on the left and right depict differences in WM saliences and GM saliences, respectively. The GM-WM pattern converged with 42 samples from each group.

ACR=anterior corona radiata; ALIC=anterior limb of the internal capsule; BCC=body of corpus callosum; CGC=cingulum (cingulate gyrus); CGH=cingulum (hippocampal portion); CP=cerebral peduncle; CR=corona radiata; CST=corticospinal tract; EC=external capsule; FX ST=fornix (cres) / stria terminalis; GCC=genu of corpus callosum; ICP=interior cerebellar peduncle; ML=medial lemniscus; MCP=middle cerebellar peduncle; PCT=pontine crossing tract; PCR=posterior corona radiata; PLIC=posterior limb of the internal capsule; PTR=posterior thalamic radiation; RLIC=retrolenticular part of the internal capsule; SCC=splenium of corpus callosum; SCR=superior corona radiata; SFO=superior fronto-occipital fasciculus; SLF=superior longitudinal fasciculus; SS=sagittal stratum; TAP=tapetum; UNC=uncinate fasciculus.

#### PLS between GM thickness and WM MD

The MB-PLS-C analyses between GM thickness and WM MD were significant (omnibus test *p* < 0.0005). The input multi-block cross-correlation matrix was decomposed into multi-block cross-correlation matrices in each latent dimension by MB-PLS-C (Figure S16). LV3 was significant and explained 11.39% of sum-of-squares covariance (Table S11).

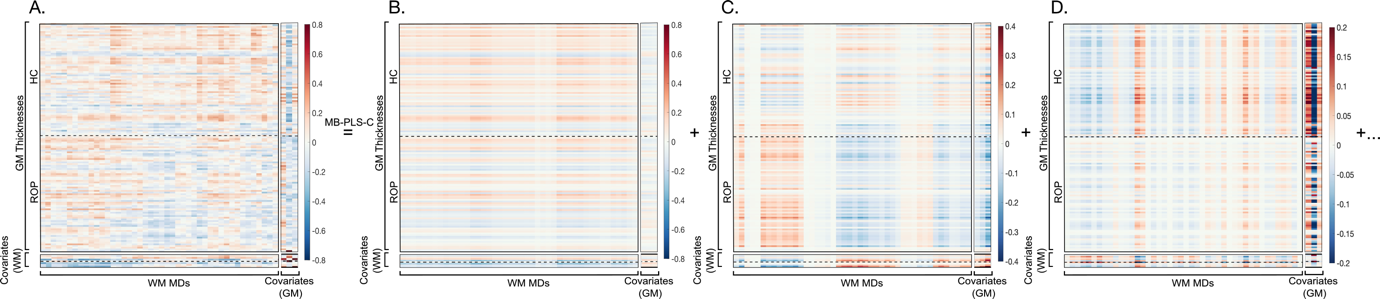

##### Figure S16. Decomposition of the input multi-block cross-correlation matrix in by MB-PLS-C between GM thickness and WM MD.

Key: A: input correlation block, B: LV1, C: LV2, D: LV3. Covariates (WM) represent age, in-scanner absolute and relative head motion, and sex by group from the top. Covariates (GM) represent intracranial volume, age, and sex from the left.

##### Table S11. MB-PLS-C latent variables contributing to the relationship between GM thickness and WM MD.

| Latent component | Contribution to 𝜎_𝑖_* (%) | | | | Sum-of-squares covariance, 𝜉𝑖* (%) | *p*-value |
| --- | --- | --- | --- | --- | --- | --- |
|  | block 1 (*𝐿_MD_^𝑇^𝐿_TH_*) | block 2 (*𝐿_MD_^𝑇^𝐿_CMD_*) | block 3 (*𝐿_CTH_^𝑇^𝐿_TH_*) | block 4 (*𝐿_CTH_^𝑇^𝐿_CMD_*) |  |  |
| LV1 | 82.57 | 14.77 | 2.04 | 0.63 | 50.15 | 0.32 |
| LV2 | 71.77 | 10.26 | 12.61 | 5.36 | 17.19 | 0.12 |
| LV3 | 25.33 | 4.27 | 54.30 | 16.10 | 11.39 | 0.001 |

Key: ^*^𝜎_𝑖_ and 𝜉_𝑖_ are the singular value and percent sum-of-squares crossblock covariance corresponding to the *i*th significant LV, 𝑖∈{1,2,3}. *L_MD_*=latent variables of MD; *L_TH_*=latent variables of thickness; *L_CMD_*=latent variables of covariates of MD; *L_CTH_*=latent variables of covariates of thickness

*Third latent variable*

LV3 accounted for 11.39% of sum-of-squares crossblock covariance (*p* = 0.001), and the data block displayed a similar correlation between the groups, mapping slightly stronger to the HC group. The covariate blocks showed negative correlations between age and GM variables and positive correlations between intracranial volume and male sex and GM variables.

Most significant saliences of GM thickness in LV3 were positively weighted and exhibited a shared GM pattern between the groups (Figure S17A). The HC group had the largest saliences in frontal and temporal regions: the right pars triangularis, pars opercularis, rostral middle frontal, caudal middle frontal, superior frontal, and middle temporal gyri. The ROP group had the largest saliences in frontal, occipital, and temporal regions and insula: bilateral lingual gyrus, left insula, middle temporal and lateral occipital gyri, and right pars triangularis. The largest saliences of WM MD were observed in the right anterior limb of the internal capsule and cingulum in the hippocampus (Figure S17B).

The HC group exhibited the most robust NSSD metric in the right rostral middle frontal, superior frontal, and caudal middle frontal gyri, pars opercularis, and pars triangularis and left precentral gyrus. The ROP group displayed the most robust NSSD metric in the left inferior temporal gyrus and insula and right rostral anterior cingulate and lingual gyri, banks of the superior temporal sulcus, and caudal anterior cingulate gyrus (Figure S17C). In the training sample (n’s as above), the HC group only showed a significant positive correlation between GM thickness and WM MD (HC: *r* = 0.35, *p* = 0.003, ROP: *r* = 0.20, *p* = 0.09). A significant correlation was not observed in the cross-validation data (n’s as above), and out-of-sample (n’s as above) didn't show a similar pattern (Figure S17D). GM and WM saliences demonstrated convergence when the sample size reached 25 per group (Figure S17E).

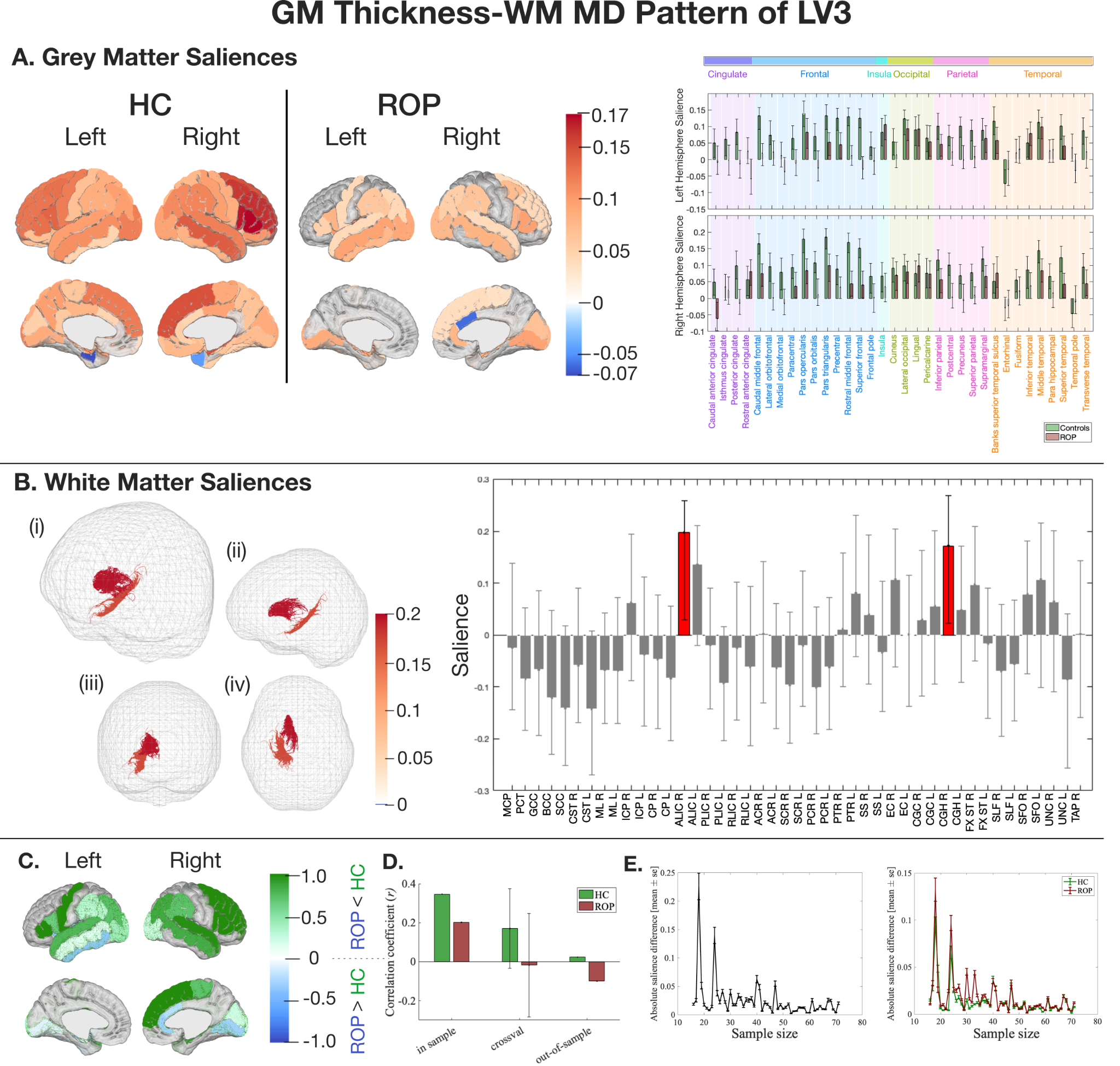

##### Figure S17. The latent pattern of LV3 between GM thickness and WM MD derived from MB-PLS-C.

Key: A) GM salience by group in HC and ROP individuals. GM saliences of HC (green) and ROP (red) with 95% confidence interval (black lines) and color-coded lobe information. B) WM saliences shared between groups. WM tracts represented in tractography are color-coded based on the salience intensity ((i) overall view (ii) from the left (iii) from the front (iv) from the bottom). The bar plot shows the salience of each WM tract with 95% confidence intervals (black lines). C) Normalised regional salience-differences. A positive score (green) shows a salience is larger in HC, and a negative score (blue) shows a salience is larger in ROP. D) A correlation coefficient between LV3 of GM and WM in the training and the test samples, and a correlation coefficient and its standard deviation from Monte Carlo cross-validation. The cross-validation data did not show a significant correlation, and out-of-sample results didn't demonstrate generalisability. E) The impact of sample size on salience intensity. Figures on the left and right depict differences in WM saliences and GM saliences, respectively. The GM-WM pattern converged with 25 samples from each group.

ACR=anterior corona radiata; ALIC=anterior limb of the internal capsule; BCC=body of corpus callosum; CGC=cingulum (cingulate gyrus); CGH=cingulum (hippocampal portion); CP=cerebral peduncle; CR=corona radiata; CST=corticospinal tract; EC=external capsule; FX ST=fornix (cres) / stria terminalis; GCC=genu of corpus callosum; ICP=interior cerebellar peduncle; ML=medial lemniscus; MCP=middle cerebellar peduncle; PCT=pontine crossing tract; PCR=posterior corona radiata; PLIC=posterior limb of the internal capsule; PTR=posterior thalamic radiation; RLIC=retrolenticular part of the internal capsule; SCC=splenium of corpus callosum; SCR=superior corona radiata; SFO=superior fronto-occipital fasciculus; SLF=superior longitudinal fasciculus; SS=sagittal stratum; TAP=tapetum; UNC=uncinate fasciculus.

#### PLS between GM surface area and WM MD

The MB-PLS-C analyses between GM surface area and WM MD were significant (omnibus test *p* < 0.0005). The input multi-block cross-correlation matrix was decomposed into multi-block cross-correlation matrices in each latent dimension by MB-PLS-C (Figure S18). LV1 was significant and explained 80.61% of sum-of-squares covariance (Table S12).

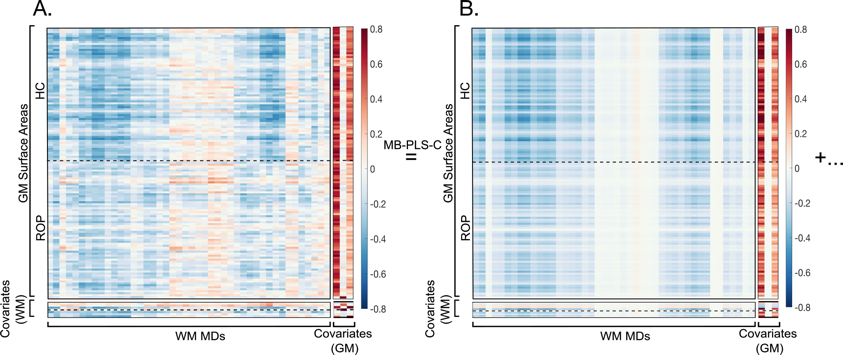

##### Figure S18. Decomposition of the input multi-block cross-correlation matrix by MB-PLS-C between GM surface area and WM MD.

Key: A: input correlation block, B: LV1. Covariates (WM) represent age, in-scanner absolute and relative head motion, and sex by group from the top. Covariates (GM) represent intracranial volume, age, and sex from the left.

##### Table S12. MB-PLS-C latent variables contributing to the relationship between GM surface area and WM MD.

| Latent component | Contribution to 𝜎_𝑖_* (%) | | | | Sum-of-squares covariance, 𝜉𝑖* (%) | *p*-value |
| --- | --- | --- | --- | --- | --- | --- |
|  | block 1 (*𝐿_MD_^𝑇^𝐿_SA_*) | block 2 (*𝐿_MD_^𝑇^𝐿_CMD_*) | block 3 (*𝐿_CSA_^𝑇^𝐿_SA_*) | block 4 (*𝐿_CSA_^𝑇^𝐿_CMD_*) |  |  |
| LV1 | 71.83 | 3.34 | 24.06 | 0.77 | 80.61 | < 0.0001 |

Key: ^*^𝜎_𝑖_ and 𝜉_𝑖_ are the singular value and percent sum-of-squares crossblock covariance corresponding to the *i*th significant LV, 𝑖∈{1}.*L_MD_*=latent variables of MD; *L_SA_*=latent variables of surface area; *L_CMD_*=latent variables of covariates of MD; *L_CSA_*=latent variables of covariates of surface area

*First latent variable*

LV1 explained 80.61% of sum-of-squares crossblock covariance (*p* < 0.0001), and both groups showed similar correlations. In the covariate blocks, total intracranial volume and male sex were positively correlated with GM surface area.

All significant saliences in LV1 were negatively weighted and exhibited a shared GM pattern between the ROP and HC groups (Figure S19A). The HC group exhibited the largest saliences in frontal and parietal regions: the bilateral lateral orbitofrontal gyrus, left superior frontal gyrus, and right precuneus, rostral middle frontal and precentral gyri. The ROP group demonstrated the largest saliences in cingulate, occipital, parietal, and temporal regions: the left inferior parietal gyrus, cuneus, and parahippocampal gyrus and right inferior temporal, inferior parietal, and posterior cingulate gyri. The largest positive saliences of WM MD were observed in the pontine crossing tract, bilateral medial lemniscus, left cingulum in the hippocampus, and right cerebral peduncle and corticospinal tract (Figure S19B).

The HC group showed the most robust NSSD metric in the bilateral superior temporal gyrus and pars orbitalis and left lateral orbitofrontal gyrus and entorhinal cortex. The ROP group exhibited the most robust NSSD metric in the bilateral lingual gyrus, left pericalcarine, cuneus, inferior parietal and lateral occipital gyri (Figure S19C). In the training sample (n’s as above), both groups showed a positive correlation between latent variables of GM surface area and WM MD (HC: *r* = 0.47, *p* < 0.001, ROP: *r* = 0.33, *p* = 0.005) and demonstrated significant correlation in the cross-validation data (n’s as above), while out-of-sample data (n’s as above) of the ROP group only demonstrated a similar correlation (Figure S19D). GM and WM saliences exhibited convergence when the sample size reached 31 per group (Figure S25E).

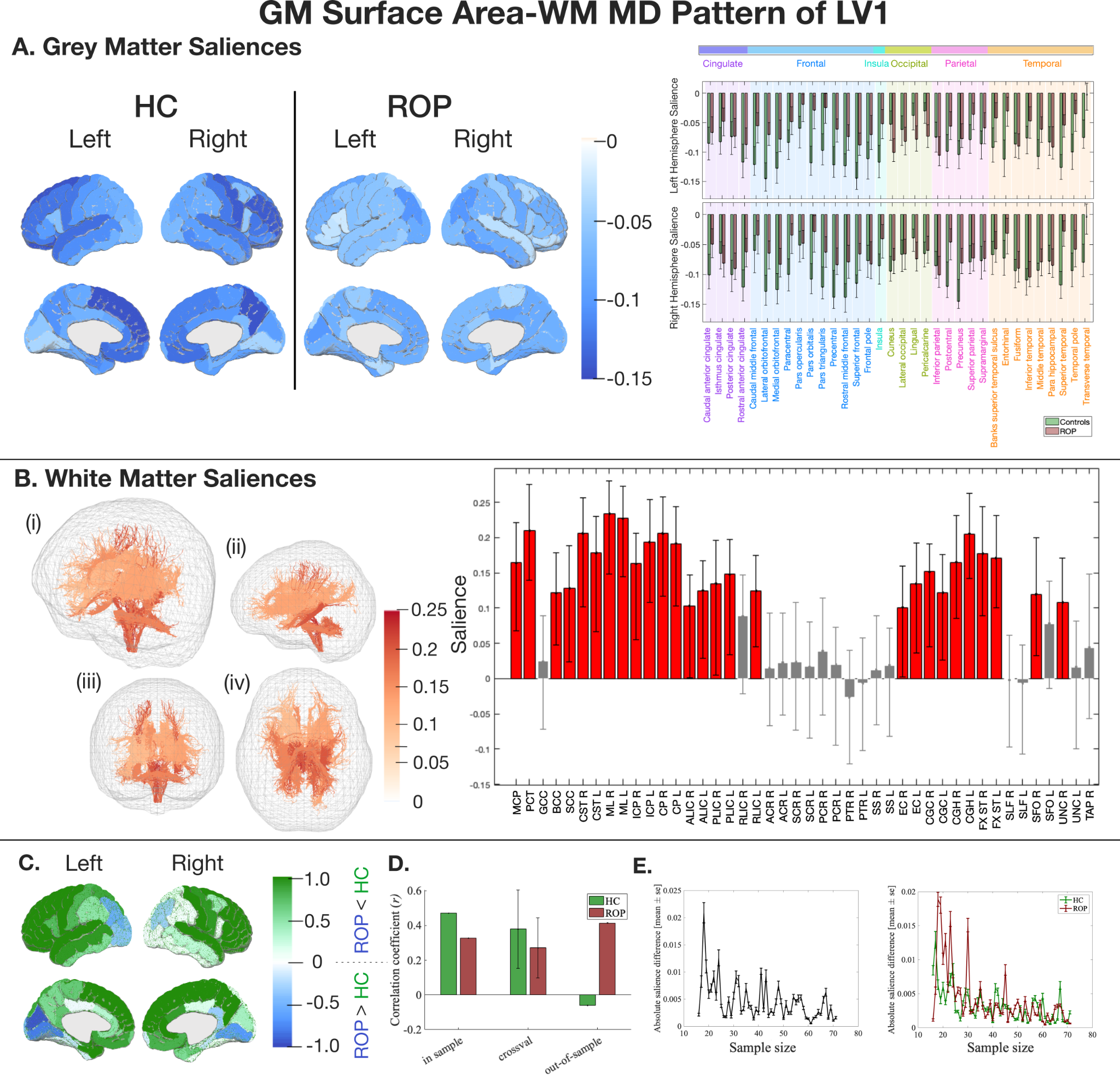

##### Figure S19. The latent pattern of LV1 between GM surface area and WM MD derived from MB-PLS-C.

Key: A) GM salience by group in HC and ROP individuals. GM saliences of HC (green) and ROP (red) with 95% confidence interval (black lines) and color-coded lobe information. All the reliable saliences were negative. B) WM saliences shared between groups. WM tracts represented in tractography are colour-coded based on the salience intensity ((i) overall view (ii) from the left (iii) from the front (iv) from the bottom). The bar plot shows the salience of each WM tract with 95% confidence intervals (black lines). C) Normalised regional salience-differences. A positive score (green) shows a salience is larger in HC, and a negative score (blue) shows a salience is larger in ROP. D) A correlation coefficient between LV1 of GM and WM in the training and the test samples, and a correlation coefficient and its standard deviation from Monte Carlo cross-validation*.* While out-of-sample data of the ROP group only showed generalisability, both groups displayed significant correlation in the cross-validation data. E) The impact of sample size on salience intensity. Figures on the left and right depict differences in WM saliences and GM saliences, respectively. The GM-WM pattern converged with 31 samples from each group.

ACR=anterior corona radiata; ALIC=anterior limb of the internal capsule; BCC=body of corpus callosum; CGC=cingulum (cingulate gyrus); CGH=cingulum (hippocampal portion); CP=cerebral peduncle; CR=corona radiata; CST=corticospinal tract; EC=external capsule; FX ST=fornix (cres) / stria terminalis; GCC=genu of corpus callosum; ICP=interior cerebellar peduncle; ML=medial lemniscus; MCP=middle cerebellar peduncle; PCT=pontine crossing tract; PCR=posterior corona radiata; PLIC=posterior limb of the internal capsule; PTR=posterior thalamic radiation; RLIC=retrolenticular part of the internal capsule; SCC=splenium of corpus callosum; SCR=superior corona radiata; SFO=superior fronto-occipital fasciculus; SLF=superior longitudinal fasciculus; SS=sagittal stratum; TAP=tapetum; UNC=uncinate fasciculus.

#### Two-group input multiblock cross-correlation Matrix, R

*GM volume and WM FA*

The input multiblock cross-correlation matrix, *R*, describes the correlations between data and covariate blocks (Figure S20). ROP individuals showed significant positive correlations between the left paracentral gyrus and bilateral superior corona radiata and between the left cerebellar cortex and left inferior cerebellar peduncle (*p* < 0.05 / ([176 rows] × [47 columns]), Bonferroni correction). Except for this, no evident correlations were found in the data block. In the covariate block, intracranial volume was significantly positively correlated with GM volumes. The correlation between male sex and GM volumes was more pronounced in the HCs than in the ROP individuals.

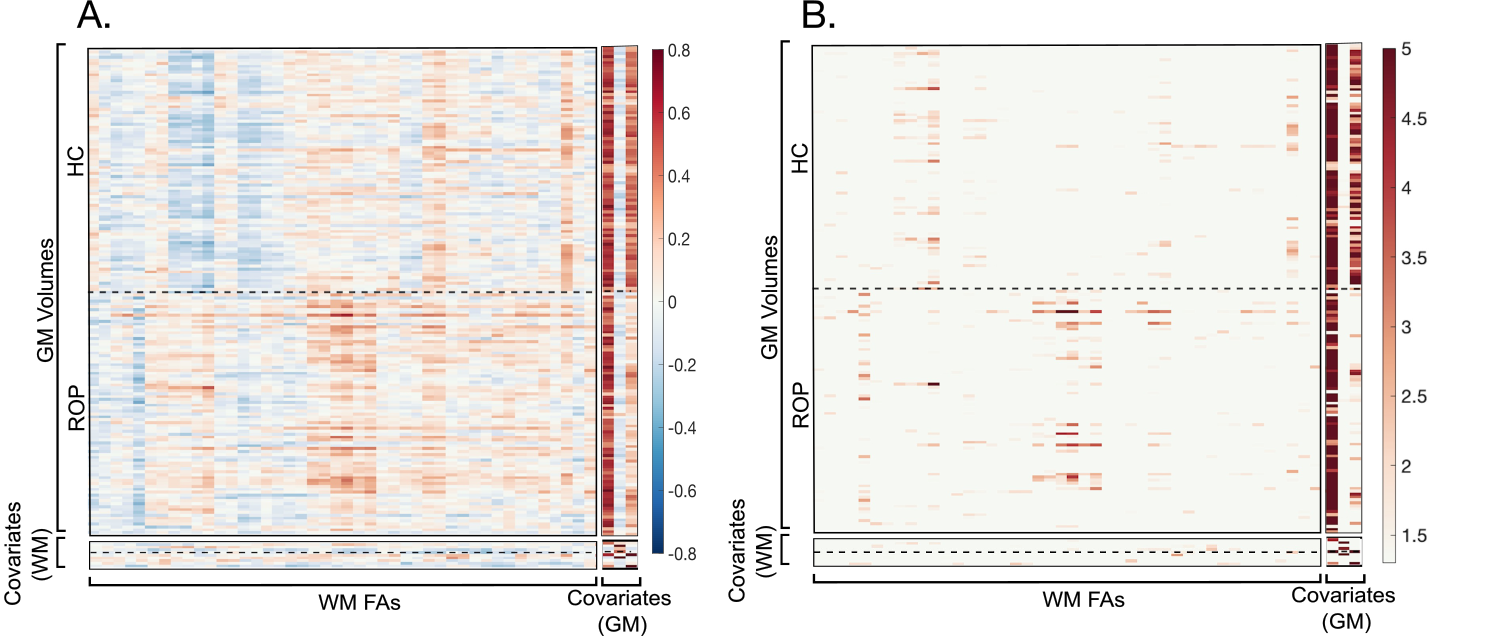

##### Figure S20. The multiblock cross-correlation matrix presenting the correlations between GM volume, WM FA, and their covariate blocks.

Key: Covariates (WM) represent age, in-scanner absolute and relative head motion, and sex by group from the top. Covariates (GM) represent intracranial volume, age, and sex from the left. A) The input matrix consists of correlation blocks between data blocks (upper left), between GM volume and their covariates (upper right), between WM FA and their covariates (lower left), and between covariates (lower right). B) Significant correlations in the input matrix are highlighted by colours (*p* < 0.05).

*GM thickness and WM FA*

Both groups didn’t show a significant correlation in the data block (*p* < 0.05 / [144 rows] × [47 columns], Bonferroni correction) (Figure S21). In the covariate block, age and GM thicknesses showed a positive correlation in controls but did not survive Bonferroni correction. There were no other obvious patterns found.

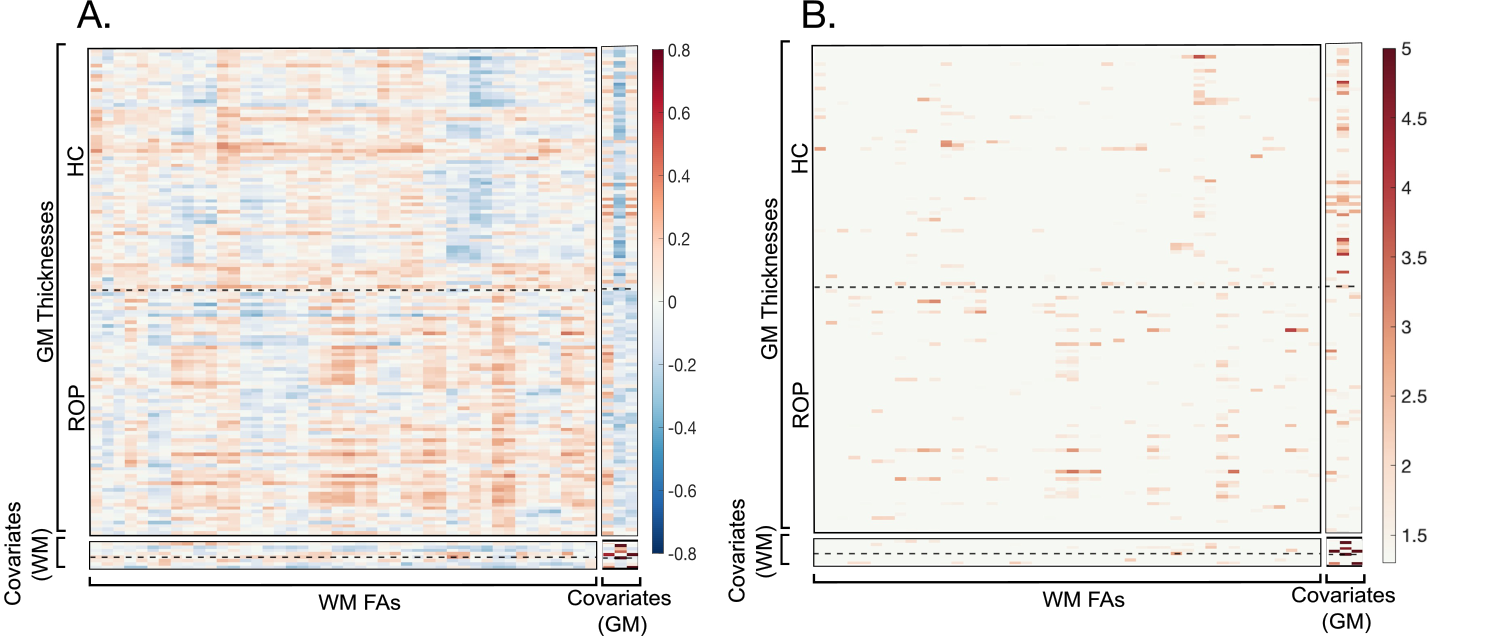

##### Figure S21. The multiblock cross-correlation matrix presenting the correlations between GM thickness, WM FA, and their covariate blocks.

Key: Covariates (WM) represent age, in-scanner absolute and relative head motion, and sex by group from the top. Covariates (GM) represent intracranial volume, age, and sex from the left. A) The input matrix consists of correlation blocks between data blocks (upper left), between GM thickness and their covariates (upper right), between WM FA and their covariates (lower left), and between covariates (lower right). B) Significant correlations in the input matrix are highlighted by colours (*p* < 0.05).

*GM surface area and WM FA*

ROP individuals showed a significant positive correlation between the left paracentral gyrus and left superior corona radiata (*p* < 0.05 / [144 rows] × [47 columns], Bonferroni correction) (Figure S22). Except for that, no evident correlations were found in the data block. In the covariate block, intracranial volume showed a significant positive correlation with GM surface area. A positive correlation between male sex and GM surface area was more pronounced in the HCs compared to the ROP individuals.

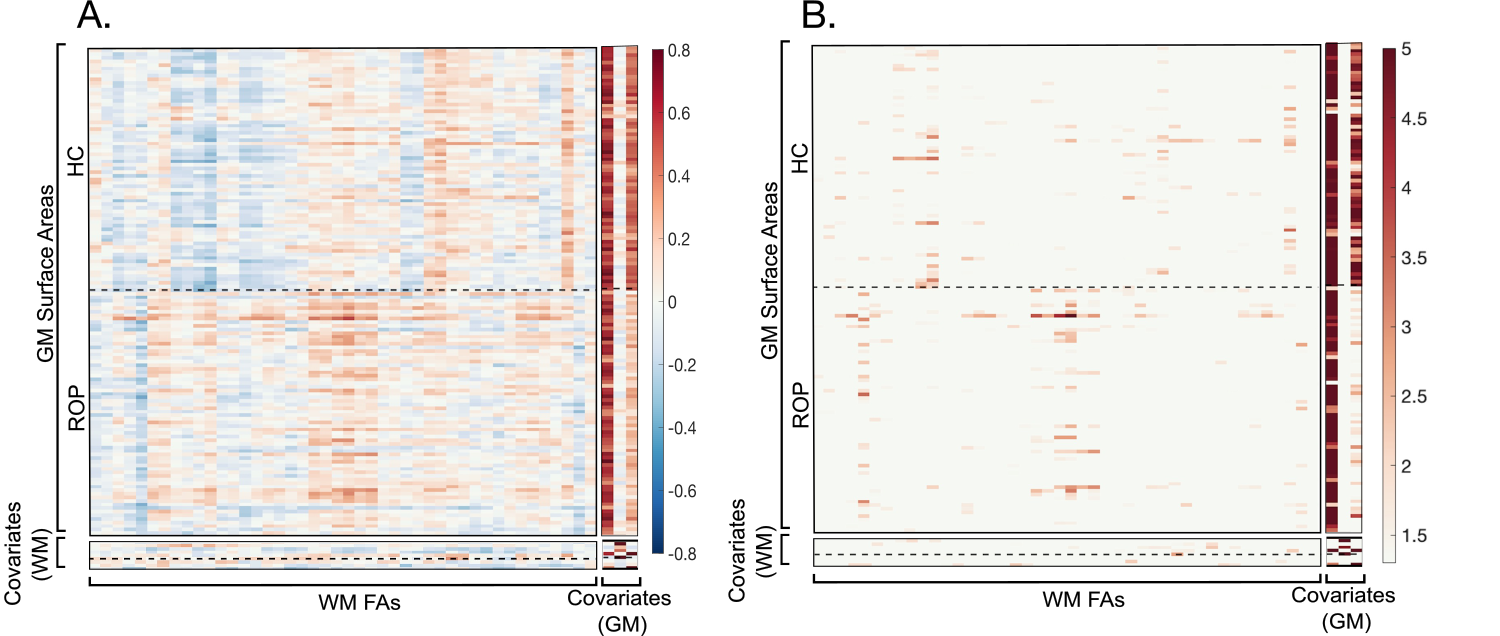

##### Figure S22. The multiblock cross-correlation matrix presenting the correlations between GM surface area, WM FA, and their covariate blocks.

Key: Covariates (WM) represent age, in-scanner absolute and relative head motion, and sex by group from the top. Covariates (GM) represent intracranial volume, age, and sex from the left. A) The input matrix consists of correlation blocks between data blocks (upper left), between GM surface area and their covariates (upper right), between WM FA and their covariates (lower left), and between covariates (lower right). B) Significant correlations in the input matrix are highlighted by colours (*p* < 0.05).

*GM volume and WM MD*

ROP individuals showed a significant negative correlation between the left nuclei accumbens and left corticospinal tract (*p* < 0.05 / [176 rows] × [47 columns], Bonferroni correction) (Figure S23). Except for it, no evident correlations were found in the data block. In the covariate block, intracranial volume showed a significant positive correlation with GM volumes. A positive correlation between male sex and GM volumes was more pronounced in the HCs compared to the ROP individuals.

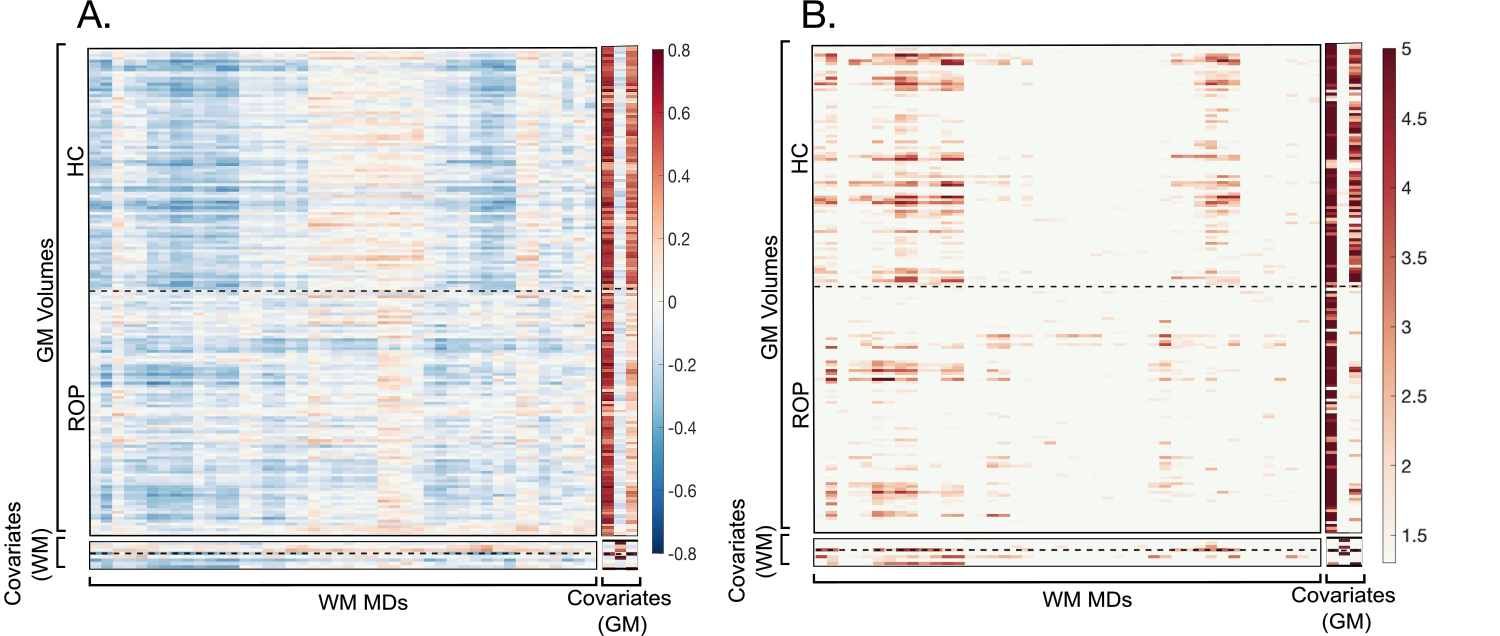

##### Figure S23. The multiblock cross-correlation matrix presenting the correlations between GM volume, WM MD, and their covariate blocks.

Key: Covariates (WM) represent age, in-scanner absolute and relative head motion, and sex by group from the top. Covariates (GM) represent intracranial volume, age, and sex from the left. A) The input matrix consists of correlation blocks between data blocks (upper left), between GM volume and their covariates (upper right), between WM MD and their covariates (lower left), and between covariates (lower right). B) Significant correlations in the input matrix are highlighted by colours (*p* < 0.05).

*GM thickness and WM MD*

Both groups didn’t show a significant correlation (*p* < 0.05 / [144 rows] × [47 columns], Bonferroni correction) (Figure S24). In the covariate block, age and GM thicknesses showed a positive correlation in controls but did not survive Bonferroni correction. There were no other obvious patterns found.

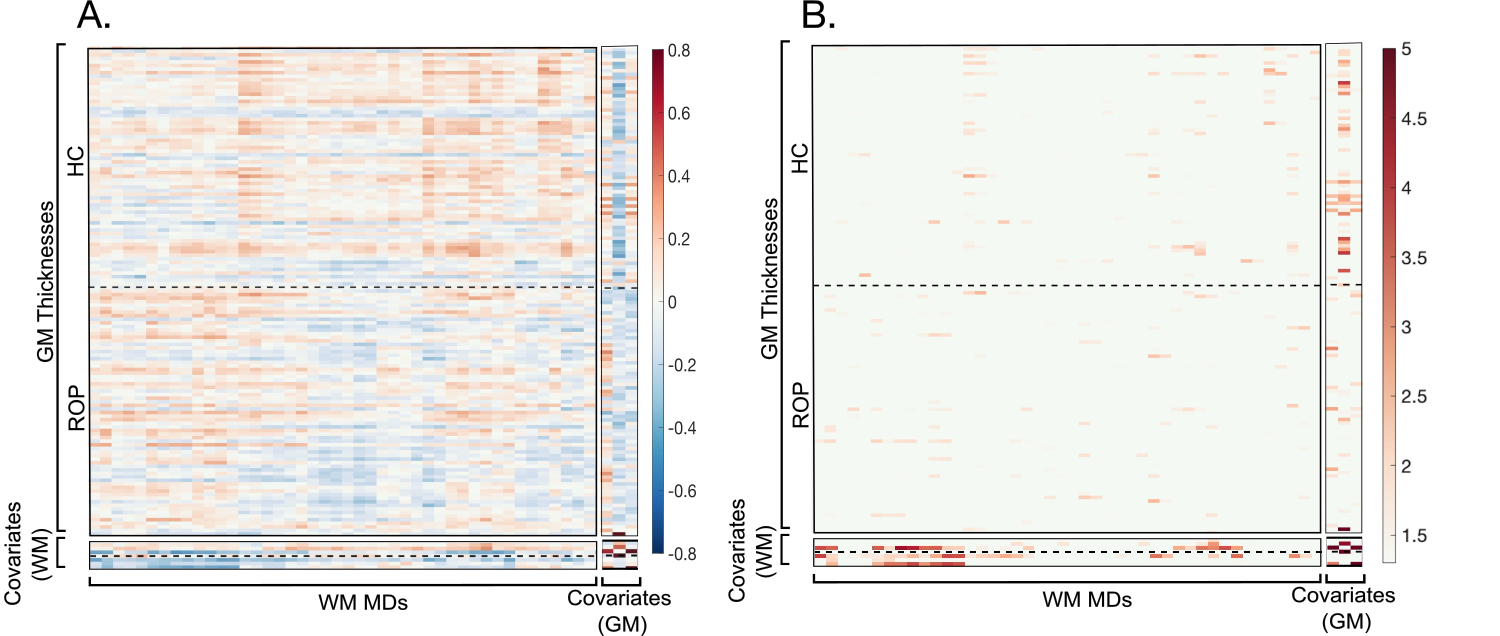

##### Figure S24. The multiblock cross-correlation matrix presenting the correlations between GM thickness, WM MD, and their covariate blocks.

Key: Covariates (WM) represent age, in-scanner absolute and relative head motion, and sex by group from the top. Covariates (GM) represent intracranial volume, age, and sex from the left. A) The input matrix consists of correlation blocks between data blocks (upper left), between GM thickness and their covariates (upper right), between WM MD and their covariates (lower left), and between covariates (lower right). B) Significant correlations in the input matrix are highlighted by colours (*p* < 0.05).

*GM surface area and WM MD*

HCs showed significant negative correlations between: the right precuneus and left medial lemniscus, left superior frontal gyrus and left cingulum in the hippocampus, left superior frontal gyrus and left medial lemniscus, left superior frontal gyrus and right medial lemniscus, left lateral orbitofrontal gyrus and right cerebral peduncle, rostral middle frontal gyrus and left cingulum in the hippocampus, left lateral orbitofrontal gyrus and right fornix stria terminalis, right precuneus and right medial lemniscus, right pars triangularis and right fornix stria terminalis, right rostral middle frontal gyrus and right medial lemniscus, right rostral middle frontal gyrus and right fornix stria terminalis, left superior frontal gyrus and right fornix stria terminalis, right precuneus and left cingulum in the hippocampus, left lateral orbitofrontal gyrus and right cerebral peduncle, and right precentral gyrus and right fornix stria terminalis (*p* < 0.05 / [144 rows] × [47 columns], Bonferroni correction) (Figure S25). In the covariate block, intracranial volume showed a significant positive correlation with GM surface areas. The correlation between male sex and GM surface areas was more pronounced in HCs compared to the ROP individuals.

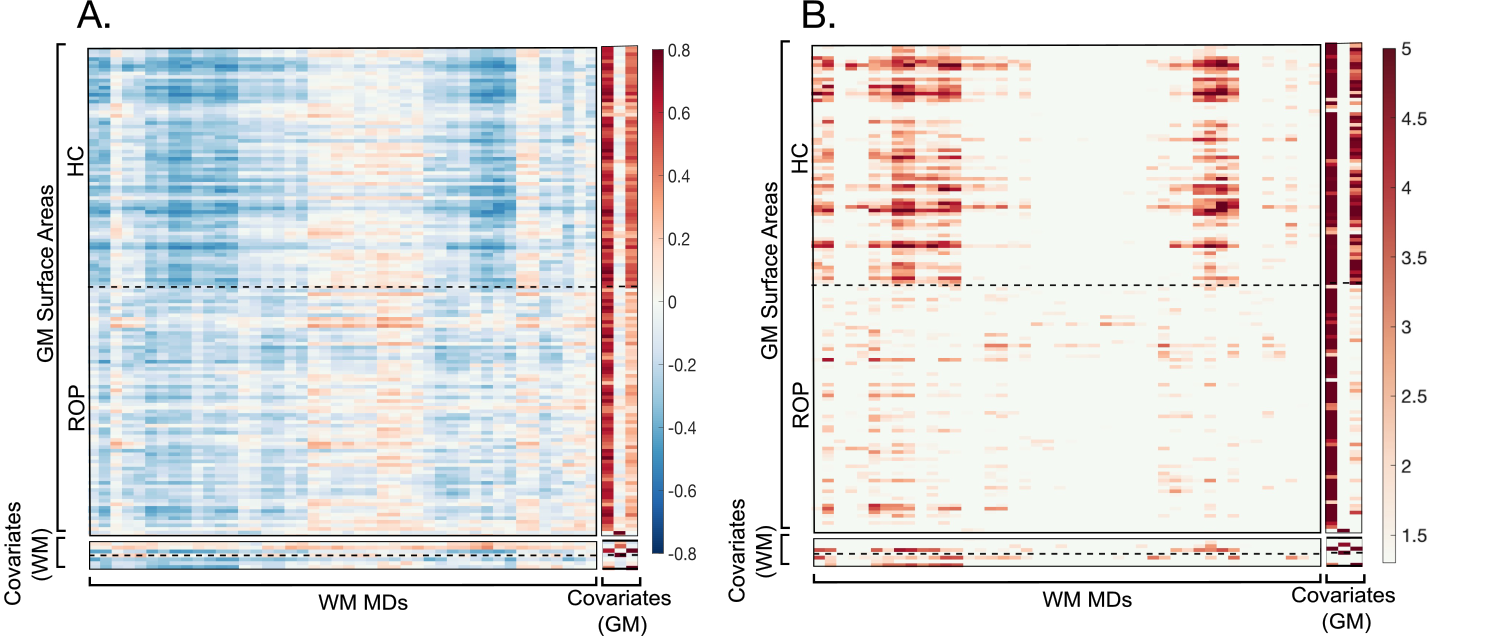

##### Figure S25. The multiblock cross-correlation matrix presenting the correlations between GM surface area, WM MD, and their covariate blocks.

Key: Covariates (WM) represent age, in-scanner absolute and relative head motion, and sex by group from the top. Covariates (GM) represent intracranial volume, age, and sex from the left. A) The input matrix consists of correlation blocks between data blocks (upper left), between GM surface area and their covariates (upper right), between WM MD and their covariates (lower left), and between covariates (lower right). B) Significant correlations in the input matrix are highlighted by colours (*p* < 0.05).

#### Correlations between cognitive abilities and latent variables of GM-WM patterns describing group differences

In the ‘GM thickness’-‘WM FA’ pattern demonstrating group differences (LV3), correlations between the GM latent variables and processing speed (ρ=0.27, p=0.022, p_adjusted_=0.06) and between the WM latent variables and episodic memory (ρ=0.26, p=0.030, p_adjusted_=0.06) and working memory (ρ=0.27, p=0.024, p_adjusted_=0.06) in the ROP group did not survive multiple comparison corrections applied using the Benjamini-Hochberg method with a 5% false discovery rate ([4 cognitive abilities] × [GM or WM latent variables]).

#### Correlations between allometric scaling maps and latent GM thickness and GM surface area patterns

Given the calculation of correlation coefficients between allometric scaling maps and latent GM patterns based on 41K vertices per hemisphere, it is important to note that even small or negligible correlations appear statistically significant due to the large number of data points. The correlation coefficients were calculated with the main direction of saliences being positive. This means that a positive correlation indicates points of the brain that are expanding during brain development have a greater salience.

*The correlation between latent GM thickness patterns and allometric scaling maps*

The ROP group showed a significant correlation between the LV2 thickness pattern and the allometric scaling map (r = 0.45, p < 1e-4), and controls also demonstrated a significant correlation (r = 0.38, p < 1e-4). Significant correlations were present in the bilateral hemisphere in the ROP (left: r = 0.59, p < 1e-4, right: r = 0.58, p < 1e-4) and HC (left: r = 0.44, p < 1e-4, right: r = 0.35, p < 1e-4) groups.

In LV3, significant correlations were observed between the GM thickness pattern and the allometric scaling maps in ROP individuals (r = -0.17, p < 1e-4) and controls (r = -0.71, p < 1e-4). The allometric scaling maps were also correlated with the GM thickness patterns in the bilateral hemisphere in the ROP (left: r = -0.15, p < 1e-4, right: r = -0.17, p < 1e-4) and HC (left: r = -0.77, p < 1e-4, right: r = -0.18, p < 1e-4) groups.

*The correlation between latent GM surface area patterns and allometric scaling maps*

The LV1 surface area pattern significantly correlated with the allometric scaling map in ROP individuals (r = 0.16, p < 1e-4), and controls also demonstrated a significant correlation (r = 0.08, p < 1e-4). Significant correlations were observed in the bilateral hemisphere in the ROP group (left: r = 0.15, p < 1e-4, right: r = 0.17, p < 1e-4) and in the left hemisphere in the HC group (r = 0.16; p < 1e-4). In the control group, the right hemisphere exhibited a statistically significant correlation with a very small effect size (r = 0.02, p < 1e-4).

Both ROP individuals (r = -0.03, p < 1e-4) and controls (r = -0.07, p < 1e-4) showed very small effect sizes in the correlations between the LV2 whole-brain GM surface area pattern and the allometric scaling maps. The allometric scaling maps had significant correlations with the bilateral hemisphere pattern of the HC group (left: r = 0.30, p < 1e-4, right: r = -0.20, p < 1e-4) and the right hemisphere pattern of the ROP group (r = -0.08, p < 1e-4), while the left hemisphere pattern of the ROP group didn't demonstrate a significant correlation (r = 0.01, p = 0.16).

Significant correlations between the allometric scaling patterns and GM thickness and GM surface area patterns in both groups indicate that these GM patterns detected by MB-PLS-C were affected by neurodevelopmental development, except for the GM surface area pattern in the right hemisphere of the HC group in LV1 and in the left hemisphere of the ROP group in LV2. The patterns showing a significant correlation with the allometric scaling patterns indicate that they are associated with neurodevelopmental changes.

1.	Lewandowski KE, Bouix S, Ongur D, Shenton ME. Neuroprogression across the Early Course of Psychosis. J Psychiatr Brain Sci. 2020;5.

12.	Mori S, Wakana S, van Zijl PCM, Nagae-Poetscher LM. MRI Atlas of Human White Matter. Elsevier; 2005.

18.	Dhollander T, Mito R, Raffelt D, Connelly A. Improved white matter response function estimation for 3-tissue constrained spherical deconvolution. 27th International Society of Magnetic Resonance in Medicine. 2019. 11 May 2019.
